# Identification of muscle weakness in older adults from normalized lower and upper limbs strength

**DOI:** 10.1101/2021.07.28.21261230

**Authors:** Pedro Pugliesi Abdalla, Lucimere Bohn, Leonardo Santos Lopes da Silva, André Pereira dos Santos, Marcio Fernando Tasinafo Junior, Ana Claudia Rossini Venturini, Anderson dos Santos Carvalho, David Martinez Gomez, Jorge Mota, Dalmo Roberto Lopes Machado

## Abstract

**Introduction:** Weakness is a natural age-related condition meaning the loss of muscle strength that impairs older adults’ mobility and quality of life. Because the relationship between muscle strength and body-size variables is non-linear, weakness is misclassified in older adults with extreme body size (e.g., light, short, heavy, or tall). This misclassification can be overcome using the allometric approach.

**Objectives:** To propose cut-off points for older adults’ weakness for upper and lower limbs muscle strength normalized by body size with the ratio standard and allometric scaling.

**Methods:** Ninety-four community-dwelling older adults (69.1% women) were assessed for 49 body-size variables (anthropometry, body composition and body indexes), handgrip strength (HGS), one maximum repetition measurement for knee extensors (1RM_knee extensors_), isokinetic knee extension peak torque at 60°/s (_knee extension_PT^60°/s^), and six-minute walk test (6MWT). Ratio standard (muscle strength/body size) and allometric scaling (muscle strength/body size^b^; when ^b^ is the allometric exponent) were applied for body-size variables that significantly were correlated with HGS, 1RM_knee extensors_ and _knee extension_PT^60°/s^. Cut-off points were computed based on ROC curve and Youden index. When there was mobility limitation (6MWT<400m) cut-off were computed according to sex.

**Results:** Absolute HGS, 1RM_knee extensors_ and _knee extension_PT^60°/s^ cut-off points were not adequate because they were associated with body size (r>0.30). But it was corrected with muscle strength normalization according to body size-variables: HGS (n=1); 1RM_knee extensors_ (n=24) and _knee extension_PT^60°/s^ (n=24). The best cut-off points, with the highest area under the curve (AUC), were found after normalization for men: HGS/forearm circumference (1.33 kg/cm, AUC=0.74), 1RM_knee extensors_/triceps skinfold (4.22 kg/mm, AUC=0.81), and _knee extension_PT^60°/s^/body mass*height^0.43^ (13.0 Nm/kg*m^0.43^, AUC=0.94); and for women: HGS/forearm circumference (1.04 kg/cm, AUC=0.70), 1RM_knee extensors_/body mass (0.54 kg/kg, AUC=0.76); and _knee extension_PT^60°/s^/body mass^0.72^ (3.14 Nm/kg^0.72^; AUC=0.82).

**Conclusion:** Normalization removes the effect of extreme body size on muscle strength and improves the accuracy to identify weakness at population level, reducing the risk of false-positive cases.

**Highlights:**

- This study proposed a new approach to identify muscle weakness in older adults based on upper and lower limbs muscle strength normalized by body size (ratio standard and allometry).
- The identification of functional limitation is more precise when procedures of muscle strength normalization is applied.

## INTRODUCTION

Muscle weakness is a natural muscle strength loss occurring along aging, and it predicts older adults’ increased risk of hospital admissions, depression, fractures and premature mortality [1-3]. Muscle weakness can predict functional disability (i.e., difficulty to perform instrumental and basic activities of daily living-ADL) like as mobility limitation [4], which is even more important than multimorbidity to forecast mortality amongst older adults [5]. As a consequence of its predictive ability, muscle weakness was used to identify geriatric syndromes such as dynapenia [6], frailty [7] and sarcopenia [8].

Muscle weakness is normally measured using muscle strength tests such as handgrip (HGS) or leg extension strength [8]. The current values to identify muscle weakness are based on absolute (non-normalized) muscle strength results [7, 9-17] or dividing absolute results by some (i.e., body mass) body-size variable (ratio standard) [18, 19]. The identification of weakness based on absolute muscle strength cut-off points may be inaccurate for lighter body mass and shorter height older adults [20-22]. In fact, the absolute values characterize lighter and shorter body size older adults as having muscle weakness, even if they sustain their instrumental and basic ADL [23]. This is a false positive muscle weakness diagnostic, that frequently leads to an unnecessarily utilization of public health resources, contributing to health burden [24]. Another topic that merits consideration is the inaccuracy of the ratio standard procedure because it overestimates the real strength of light/short older adults and underestimates it for tall/heavy ones [23]. These limitations are a consequence of the nonlinear relationship between muscle strength and body-size variables [20-22]. To overcome these constraints, the utilization of allometric scaling, that contemplates power and sensitivity in the nonlinear relationship between muscle strength and body size with the allometric exponent (^b^) might represent an adequate option [20-23].

Previous studies reported already the power function ratio in older adults between HGS and body-size variables as body mass (^b=0.63^ or ^0.40^ or ^0.31^) [20-22], height (^b=1.84^) [21] and fat-free mass (FFM) (^b=0.46^) [21] and between leg extension strength and body mass (^b=0.67^ or ^0.69^ or ^0.72^ or ^0.74^ or ^0.96^) [23, 25]. Indeed, scaling HGS by body size (example: HGS/heigth^1.84^) removes the effect of body size on muscle strength [21], but the scaling muscle strength by body size to determine muscle weakness cut-off points has not been considered from HGS and knee extension in isokinetic dynamometer, excepting the one maximum repetition measurement for knee extensors (1RM_knee extensors_) scaled to body mass [23]. Besides, important body-size variables related to mobility and ADL (e.g. fat mas [26], FFM [27] and leg length [28]) were not utilized to scaling muscle strength and create muscle weakness cut-off points.

Thus, our objective is to propose cut-off points for older adults’ weakness with upper and lower limbs muscle strength normalized by body-size with the ratio standard and allometric scaling. We hypothesize that the normalization of muscle strength by ratio standard and allometry can be a way to approach muscle strength regardless of body size, which should reduce the risk of bias in identifying false-positive cases of vulnerable older people.

## MATERIAL AND METHODS

### Design and Study population

This is a cross-sectional study conducted from October 2016 to May 2017 at the University Hospital of Ribeirao Preto School of Medicine, University of São Paulo, Brazil (HC-FMRP-USP). The study was approved by the HC-FMRP-USP institutional review board. Older adults were voluntarily recruited and assigned an informed consent. This manuscript followed the guidelines from The Strengthening the Reporting of Observational Studies in Epidemiology (STROBE) conference list, and the completed checklist is attached.

The sample consisted of 94 community-dwelling older adults (>60 years old, 69.1% women) recruited in projects for older adults of USP and in health community services. Inclusion criteria were walk independently, absent limitation to execute all procedures, acute infections, cancer diagnosis, hip or knee prostheses, unstable cardiovascular condition, stroke sequelae, tumors, and weight loss >3 kg in the last three months. The exclusion criteria were discontinuity in the study and cognition impairment.

A sample size calculation (n=[ZySD/ε]^2^) with trust level (Zy=0,95), greater compatible population variability founded in the literature (SD of 1RM_knee extensors_: ±19.96 kg) [29, 30] and maximum desired error (ε≤8.0 kg) was performed and identified a minimum sample size of n=24 for each sex.

### Procedures

A multidisciplinary health team (nurses, nutritionists, pharmacists, physical education professors, physicians, and physiotherapists) performed data collection. The appraisers were the same in each test. Data collection occurred on three non-consecutive days: 1^st^) recruitment: inclusion criteria verification and cognition assessment; by phone calls (inclusion criteria) or face to face (cognition assessment) of older adults who agreed to participate in the study; 2^nd^) anthropometrics, body composition, HGS, mobility and physical activity level assessment; and 3^rd^) lower limbs muscle strength assessment.

### Cognition Assessment

The validated Mini Mental State Examination (MMSE) was used to assess participants’ cognition status [31]. Those who have MMSE≤12 were excluded [32].

### Measure of Body-Size Variables

Forty-nine body-size variables (SUPPLEMENT A) were collected to propose allometric exponents and to normalize performance in muscle strength tests. The selection of these variables were based on those previously used to calculate body indices [33-41], and involved anthropometric measurements [42] and body composition (Dual Energy X-ray Absorptiometry-DXA and bioelectrical impedance analysis-BIA), as briefly detailed below.

### Body Indexes

The body indexes derived from anthropometry were body mass index (BMI, kg/m^2^) [33], body mass*height [34], human body surface area (SA, m^2^) [35], absolute mid-arm muscle circumference (MAMC, cm) [36], corrected arm muscle area (CAMA, cm) [37], arm fat area (AFA,cm^2^) [38], FFM [39] and fat mass (obtained by body mass difference). The body indexes derived from body composition were lean soft tissue (LST) of arms and legs, appendicular skeletal muscle mass (ASM), ASM/height (m)^2^ [40], FFM (estimated from BIA) [41] and DXA, when fat mass were estimated by body mass difference.

### Mobility Measurement

The cut-off points for muscle weakness were established based on mobility limitation. Mobility performance was verified based on the six-minute walk test (6MWT) carried out in a corridor 30-meter length. Along this path, at every three meters there was a cone to help researcher to precisely identify the walked distance [28]. Participants were instructed to cover the longest distance walking as faster as they could during the six-minute time. Nevertheless, participants could slow down, interrupt the walking, and resume the test whenever desired, although time was not paused. Total walked distance was recorded and mobility disability was characterized when the 6MWT<400 m [43].

### Muscle Strength Measurements

Muscle strength was measured using HGS, one maximum repetition measurement for knee extensors (1RM_knee extensors_) and isokinetic knee extension peak torque at a velocity of 60°/s (_knee extension_PT^60°/s^). The maximum HGS was measured with a manual dynamometer (Jamar®, model 5030J1) using a previously published protocol [44]. Three attempts were performed, one minute apart, with the dominant hand and the highest result was recorded in kg as HGS [45, 46]. The 1RM_knee extensors_ was estimated in a leg extension machine (Lion Fitness® model LFS) with a submaximal repetition protocol: 1RM=weight lifted/(1.0278-[0.0278*n° of reps]) [47]. The detailed protocol was published elsewhere [23]. The isokinetic knee extension peak torque at 60°/s (_knee extension_PT^60°/s^) of the right lower limb was recorded with the Biodex (model System 4 Pro) isokinetic dynamometer and results are in newton-meter (Nm) according to standardized protocol [48].

### Physical Activity Level Measurement

The International Physical Activity Questionnaire - Short Version was used to get physical activity level [49]. Physical activity level was dichotomized into sedentary (0) and irregularly active, active or very active (1). These two categories were introduced in the models to provide allometric exponents.

### Muscle Strength Normalization Procedures (Ratio Standard and Allometric Scaling)

HGS, 1RM_knee extensors_ and _knee extension_PT^60°/s^ were considered in three different ways: 1) absolute (non-normalized); 2) ratio standard (muscle strength/body-size variable); and 3) allometrically adjusted (muscle strength/body-size variable^b^).

Allometric exponents (^b^) were proposed only for body-size variables that showed significant correlation (Pearson’s correlation) with muscle strength. To generate the allometric exponents, muscle strength (Y) and body-size variables (X) were converted to natural logarithm (ln) and the slope of regression line is allometric exponent (^b^), according to more detail previously published [21]. Therefore, allometric exponents were discarded when the interaction (ln body-size variable*age*sex*physical activity level) was significant or when there was multicollinearity in the linear regression (variance inflation factor [VIF]>10) [50].

We also consider other allometric exponents (^b^) of the literature, as described in Table 1.

**Table 1.** Allometric exponents (^b^) proposed in previous studies.

| Authors | Normalized muscle strength for body-size variable |
| --- | --- |
| Jaric et al., (2003) [58] | General muscle strength/body mass<sup>0.67</sup> |
| Foley et al., (1999) [20] | HGS/body mass<sup>0.40</sup> |
| Pua (2006) [22] | HGS/body mass<sup>0.63</sup> |
| Maranhão Neto et al., (2017) [21] | HGS/body mass<sup>0.31</sup><br>HGS/height<sup>1.84</sup> |
| Abdalla et al., 2020 [23] | 1RM<sub>knee extensors</sub>/body mass<sup>0.69</sup><br>1RM<sub>knee extensors</sub>/body mass<sup>0.96</sup> |
| Davies and Dalsky (1997) [25] | knee extensionPT<sup>60°/s</sup>/body mass<sup>0.67</sup><br>knee extensionPT<sup>60°/s</sup>/body mass<sup>0.72</sup><br>knee extensionPT<sup>60°/s</sup>/body mass<sup>0.74</sup> |
| Segal et al., (2008) [34] | knee extensionPT<sup>60°/s</sup>/body mass*height<sup>0.97</sup> |

HGS=handgrip strength; 1RM<sub>knee extensors</sub>=one maximum repetition measurement for knee extensors; knee extensionPT<sup>60°/s</sup>=isokinetic knee extension peak torque at 60°/s

In order to verify whether normalization removed the influence of body size on muscle strength, the correlation between normalized muscle strength and body-size variables (body mass, height and body-size used) should be negligible (r≤30) [51].

### Statistical Analysis

#### Proposition of Cut-off Points for Muscle Weakness

Absolute muscle strength and normalized by ratio standard or allometric scaling had their area under the curve (AUC) quantified by the ROC curve. The Youden index [52] selected the most appropriate cut-off points with the best relationship between sensitivity and specificity for the primary outcome (functional limitation: 6MWD<400) [43].

The cut-off points were considered adequate when they have AUC≥0.70 [53] simultaneously for both sexes (p<0.05) and when the correlation between muscle strength and body-size variables (body mass, height and body-size used) were negligible (r≤0.30) [51].

For each muscle strength test (HGS, 1RM_knee extensors_ and _knee extension_PT^60°/s^), way (non-normalized, ratio standard and allometric scaling) and for each and sex was selected the adequate cut-off point according the superior accuracy. When there was a tie in accuracy, the variable with the greatest sensitivity or specificity was chosen. Finally, the AUC - ROC curves of non-normalized and normalized muscle strength were compared with each other to decide the best cut-off point.

The analyzes were performed using the SPSS 25.0 statistical package, and the ROC curves and Youden index in MedCalc 15.2 with a previously established level of significance (α=5%).

## RESULTS

Sample was encompassed by 100 older adults (69 women) who agreed to participate in the study. From those, 6 were excluded for different reasons, as the stages of the study proceeded, as detailed in Figure 1. Therefore, the final sample comprised 29 older men (31%) and 65 older women (69%).

**Figure 1.**
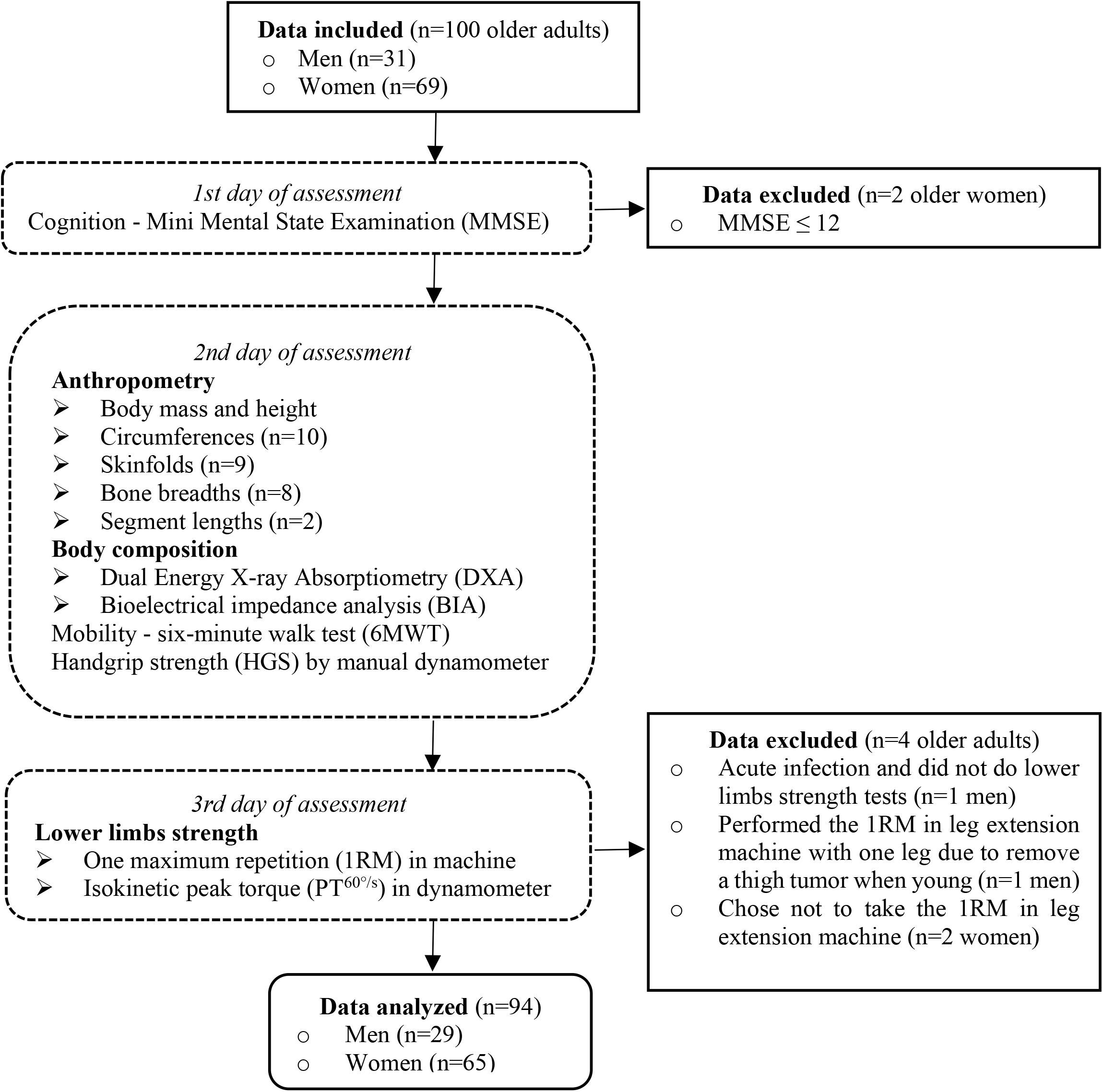
Study phases and data from older adults included, excluded, analyzed and procedure flow.

Sample characterization according to sex is shown in Table 2. Twenty-five women (38.5%) and seven men (24.1%) had functional limitation (6MWT<400 m).

**Table 2.** Descriptive analysis and significant correlations of muscle strength with body-size variables in older men and women (n=94).

| Variables | Older Men (n=29) |  |  |  | Older Women (n=65) |  |  |  | Correlation (r) with Muscle Strength |  |  |
| --- | --- | --- | --- | --- | --- | --- | --- | --- | --- | --- | --- |
|  | M | 95% CI |  | SD | M | 95% CI |  | SD | HGS (kg) | Knee Extension |  |
|  |  | LL | UL |  |  | LL | UL |  |  | 1RM (kg) | PT<sup>60°/s</sup> (Nm) |
| Age (years) | 71.2 | 68.5 | 73.9 | 7.1 | 69.7 | 68.2 | 71.2 | 6.1 |  |  |  |
| Mini-Mental State Examination (0-19) | 17.6 | 16.9 | 18.2 | 1.8 | 17.4 | 16.9 | 17.8 | 1.8 |  |  |  |
| Body-size variables |  |  |  |  |  |  |  |  |  |  |  |
| Anthropometry |  |  |  |  |  |  |  |  |  |  |  |
| Body mass (kg) | 73.0 | 67.7 | 78.3 | 13.9 | 66.9 | 64.0 | 69.8 | 11.6 | 0.37<sup>†</sup> | 0.39<sup>†</sup> | 0.40<sup>†</sup> |
| Height (m) | 1.7 | 1.6 | 1.7 | 0.1 | 1.6 | 1.5 | 1.6 | 0.1 | 0.71<sup>†</sup> | 0.62<sup>†</sup> | 0.68<sup>†</sup> |
| Circumferences (cm) |  |  |  |  |  |  |  |  |  |  |  |
| Forearm | 26.0 | 25.2 | 26.8 | 2.0 | 23.8 | 23.3 | 24.4 | 2.2 | 0.50<sup>†</sup> | 0.40<sup>†</sup> | 0.46<sup>†</sup> |
| Calf | 35.8 | 34.4 | 37.1 | 3.5 | 34.8 | 34.1 | 35.5 | 2.9 | 0.34<sup>*</sup> | 0.37<sup>†</sup> | 0.37<sup>†</sup> |
| Chest | 97.8 | 94.3 | 101.4 | 9.4 | 93.2 | 91.3 | 95.1 | 7.7 | 0.26<sup>*</sup> | 0.32<sup>*</sup> | 0.36<sup>†</sup> |
| Waist | 92.1 | 87.8 | 96.5 | 11.4 | 86.5 | 84.0 | 89.0 | 10.0 |  | 0.26<sup>*</sup> | 0.30<sup>*</sup> |
| Skinfold thickness (mm) |  |  |  |  |  |  |  |  |  |  |  |
| Triceps | 15.2 | 12.9 | 17.5 | 6.0 | 25.8 | 24.1 | 27.4 | 6.7 | -0.39<sup>†</sup> | -0.22<sup>*</sup> | -0.25<sup>*</sup> |
| Biceps | 8.0 | 6.7 | 9.4 | 3.5 | 15.4 | 14.0 | 16.7 | 5.4 | -0.40<sup>†</sup> | -0.28<sup>*</sup> | -0.31<sup>*</sup> |
| Midaxillary | 18.5 | 15.5 | 21.5 | 7.8 | 23.9 | 22.2 | 25.6 | 6.9 | -0.26<sup>*</sup> |  |  |
| Pectoral | 16.8 | 14.7 | 18.9 | 5.5 | 14.6 | 13.0 | 16.2 | 6.4 | 0.21<sup>*</sup> |  |  |
| Suprailiac | 19.7 | 15.8 | 23.5 | 10.1 | 29.7 | 27.8 | 31.7 | 7.8 | -0.29<sup>*</sup> |  |  |
| Abdominal (vertical) | 26.2 | 23.1 | 29.3 | 8.1 | 33.7 | 31.5 | 35.9 | 8.8 | -0.22<sup>*</sup> |  |  |
| Thigh (midline) | 17.9 | 15.0 | 20.8 | 7.6 | 32.1 | 29.5 | 34.8 | 10.7 | -0.35<sup>*</sup> | -0.27<sup>*</sup> | -0.38<sup>†</sup> |
| Medial calf | 11.8 | 9.3 | 14.3 | 6.6 | 23.8 | 21.9 | 25.7 | 7.6 | -0.41<sup>†</sup> | -0.28<sup>*</sup> | -0.33<sup>*</sup> |
| Bone breadths (mm) |  |  |  |  |  |  |  |  |  |  |  |
| Biacromial | 39.9 | 38.9 | 41.0 | 2.8 | 37.1 | 36.6 | 37.6 | 2.1 | 0.63<sup>†</sup> | 0.58<sup>†</sup> | 0.60<sup>†</sup> |
| Bitrochanteric | 33.7 | 33.1 | 34.3 | 1.7 | 33.4 | 32.8 | 34.0 | 2.3 |  | 0.20<sup>*</sup> | 0.22<sup>*</sup> |
| Ankle (bimalleolar) | 7.0 | 6.8 | 7.2 | 0.5 | 6.3 | 6.2 | 6.4 | 0.4 | 0.59<sup>†</sup> | 0.47<sup>†</sup> | 0.52<sup>†</sup> |
| Elbow | 6.7 | 6.5 | 6.9 | 0.5 | 5.8 | 5.7 | 6.0 | 0.5 | 0.53<sup>†</sup> | 0.40<sup>†</sup> | 0.44<sup>†</sup> |
| Wrist | 5.7 | 5.6 | 5.9 | 0.4 | 5.1 | 5.0 | 5.1 | 0.4 | 0.52<sup>†</sup> | 0.37<sup>†</sup> | 0.44<sup>†</sup> |
| Chest | 30.9 | 29.9 | 31.9 | 2.6 | 27.8 | 27.4 | 28.3 | 1.8 | 0.58<sup>†</sup> | 0.49<sup>†</sup> | 0.59<sup>†</sup> |
| Segment lengths (cm) |  |  |  |  |  |  |  |  |  |  |  |
| Knee height | 53.5 | 52.4 | 54.5 | 2.7 | 49.5 | 49.0 | 50.0 | 2.1 | 0.62<sup>†</sup> | 0.55<sup>†</sup> | 0.58<sup>†</sup> |
| Half arm span | 87.3 | 85.5 | 89.2 | 4.8 | 80.8 | 79.8 | 81.7 | 3.8 | 0.71<sup>†</sup> | 0.62<sup>†</sup> | 0.58<sup>†</sup> |
| Body indexes |  |  |  |  |  |  |  |  |  |  |  |
| Derived from anthropometry |  |  |  |  |  |  |  |  |  |  |  |
| Body mass*height (kg*m) | 123.3 | 112.6 | 134.1 | 28.2 | 104.7 | 99.8 | 109.7 | 20.0 | 0.49<sup>†</sup> | 0.49<sup>†</sup> | 0.52<sup>†</sup> |
| SA (m²) | 1.9 | 1.8 | 1.9 | 0.2 | 1.7 | 1.7 | 1.8 | 0.2 | 0.47<sup>†</sup> | 0.48<sup>†</sup> | 0.50<sup>†</sup> |
| MAMC (cm) | 24.2 | 23.0 | 25.3 | 2.9 | 21.9 | 21.3 | 22.5 | 2.6 | 0.45<sup>†</sup> | 0.39<sup>†</sup> | 0.45<sup>†</sup> |
| CAMA (cm²) | 37.1 | 32.7 | 41.6 | 11.7 | 32.2 | 29.9 | 34.5 | 9.2 | 0.37<sup>†</sup> | 0.33<sup>†</sup> | 0.40<sup>†</sup> |
| AFA (cm²) | 16.3 | 14.0 | 18.6 | 6.0 | 22.9 | 21.3 | 24.6 | 6.6 | -0.23<sup>*</sup> |  |  |
| FFM<sub>(LEAN et al., 1996)</sub> (kg) | 52.1 | 49.6 | 54.6 | 6.6 | 37.1 | 36.0 | 38.2 | 4.6 | 0.75<sup>†</sup> | 0.66<sup>†</sup> | 0.67<sup>†</sup> |
| Fat mass<sub>(LEAN et al., 1996)</sub> (kg) | 20.9 | 17.7 | 24.0 | 8.2 | 29.8 | 27.9 | 31.7 | 7.8 | -0.22<sup>*</sup> |  |  |
| Derived from body composition |  |  |  |  |  |  |  |  |  |  |  |
| Left arm LST (kg) | 2.4 | 2.1 | 2.6 | 0.6 | 1.5 | 1.4 | 1.6 | 0.3 | 0.72<sup>†</sup> | 0.62<sup>†</sup> | 0.66<sup>†</sup> |
| Right arm LST (kg) | 2.8 | 2.5 | 3.0 | 0.6 | 1.8 | 1.7 | 1.9 | 0.4 | 0.72<sup>†</sup> | 0.61<sup>†</sup> | 0.60<sup>†</sup> |
| Left leg LST (kg) | 7.7 | 7.1 | 8.3 | 1.6 | 5.5 | 5.3 | 5.8 | 1.0 | 0.67<sup>†</sup> | 0.64<sup>†</sup> | 0.64<sup>†</sup> |
| Right leg LST (kg) | 8.0 | 7.4 | 8.6 | 1.6 | 5.7 | 5.4 | 5.9 | 1.0 | 0.70<sup>†</sup> | 0.64<sup>†</sup> | 0.65<sup>†</sup> |
| Arms LST (kg) | 5.1 | 4.7 | 5.6 | 1.2 | 3.3 | 3.2 | 3.5 | 0.7 | 0.74<sup>†</sup> | 0.63<sup>†</sup> | 0.64<sup>†</sup> |
| Legs LST (kg) | 15.7 | 14.6 | 16.9 | 3.1 | 11.2 | 10.7 | 11.6 | 1.9 | 0.69<sup>†</sup> | 0.65<sup>†</sup> | 0.66<sup>†</sup> |
| ASM (kg) | 20.9 | 19.3 | 22.5 | 4.2 | 14.5 | 13.9 | 15.1 | 2.5 | 0.72<sup>†</sup> | 0.65<sup>†</sup> | 0.66<sup>†</sup> |
| ASM/height² (kg/m²) | 7.3 | 7.0 | 7.7 | 1.0 | 6.0 | 5.7 | 6.2 | 0.9 | 0.55<sup>†</sup> | 0.51<sup>†</sup> | 0.48<sup>†</sup> |
| FFM<sub>(BAUMGARTNER et al., 1991)</sub> (kg) | 54.3 | 51.3 | 57.3 | 7.7 | 45.5 | 44.0 | 47.0 | 6.1 | 0.60<sup>†</sup> | 0.55<sup>†</sup> | 0.57<sup>†</sup> |
| FFM<sub>(DXA)</sub> (kg) | 51.5 | 47.9 | 55.0 | 9.4 | 38.8 | 37.4 | 40.3 | 5.8 | 0.68<sup>†</sup> | 0.59<sup>†</sup> | 0.61<sup>†</sup> |
| Fat mass<sub>(DXA)</sub> (kg) | 21.5 | 18.8 | 24.2 | 7.1 | 28.1 | 26.3 | 29.9 | 7.2 | -0.20<sup>*</sup> |  |  |
| Mobility |  |  |  |  |  |  |  |  |  |  |  |
| Six-minute walk test (6MWT) | 464.7 | 431.1 | 498.3 | 88.3 | 412.7 | 389.9 | 435.5 | 92.0 |  |  |  |
| Functional limitation (6MWT<400m); % |  | 24.1% |  |  |  | 38.5% |  |  |  |  |  |
| Muscle strength |  |  |  |  |  |  |  |  |  |  |  |
| HGS (kg) | 36.4 | 33.1 | 39.7 | 8.6 | 24.1 | 23.0 | 25.2 | 4.5 |  |  |  |
| 1RM<sub>knee extensors</sub> (kg) | 66.8 | 56.9 | 76.7 | 26.0 | 40.8 | 36.9 | 44.8 | 15.9 |  |  |  |
| n° of reps to estimate 1RM | 7.2 | 6.3 | 8.1 | 2.3 | 6.6 | 6.0 | 7.2 | 2.4 |  |  |  |
| knee extension PT<sup>60°/s</sup> (Nm) | 119.8 | 102.4 | 137.2 | 45.6 | 73.2 | 66.8 | 79.6 | 25.9 |  |  |  |

\*p<0.05 and †p<0.001 (statistically significant correlation).
Note: M=mean; CI=confidence interval; LL=lower limit; UL=upper limit; SD=standard deviation; HGS=handgrip strength; 1RM=one maximum repetition; 1RM<sub>knee extensors</sub>=one maximum repetition measurement for knee extensors; PT<sup>60°/s</sup>=isokinetic peak torque at 60°/s; <sub>knee extension</sub>PT<sup>60°/s</sup>=isokinetic knee extension peak torque at 60°/s; Nm=Newton meter; SA=human body surface area; MAMC=mid-arm muscle circumference; CAMA=corrected arm muscle area; AFA=arm fat area; FFM=fat-free mass; LST=lean soft tissue; ASM=appendicular skeletal muscle mass; DXA=Dual-energy X-ray absorptiometry.

The correlations between muscle strength and body-size variables are also shown in Table 2. Most of the body-size variables showed a significant correlation with muscle strength (r=-0.41 to 0.75; p<0.05).

Allometric exponents were proposed for those body-size variables that showed a significant relationship with muscle strength (Table 2). Linear regressions to obtain allometric exponents are shown in SUPPLEMENT B. All regressions were significant to explain muscle strength (p<0.05), with adjusted R^2^ ranging from 0.39 to 0.61. The regression coefficients (β) obtained for each body-size variable represent the allometric exponents obtained. For HGS, the allometric exponents of triceps, pectoral, abdominal and thigh skinfolds were discarded because the interaction terms were statistically significant (p<0.05) and have accentuated multicollinearity (VIF>10). The remaining allometric exponents were used to perform normalization (for example, 1RM_knee extensors_/body mass^0.44^).

The sex-specific cut-off points proposed for HGS, 1RM_knee extensors_ and _knee extension_PT^60°/s^ (non-normalized, ratio standard and allometric scaling) to identify muscle weakness are presented in the SUPPLEMENT C. In the same supplement there are also presented correlations between muscle strength and body size (body mass, height and body-size variable used in normalization).

Non-normalized HGS, 1RM_knee extensors_ and _knee extension_PT^60°/s^ cut-off points to identify muscle weakness were not adequate for both sexes or because they did not present AUC≥0.70 (p<0.05) or because they had a significant association with body size (r>0.30; p<0.05) (SUPPLEMENT C).

Table 3 shows the cut-off points based on the ratio standard and allometric scaling classified as adequate.

**Table 3.** Adequate cut-off points (AUC≥0.70 simultaneously in both sexes and r≤0.30 with body size) of handgrip strength (HGS), one maximum repetition measurement for knee extensors (1RM_knee extensors_) and isokinetic knee extension peak torque at 60°/s (_knee extension_PT^60°/s^) to identify muscle weakness.

| Variable | Unit | Men (n=29) |  |  |  | Women (n=65) |  |  |  |
| --- | --- | --- | --- | --- | --- | --- | --- | --- | --- |
|  |  | AUC | Cut-off point<br>(≤) | Sens<br>(%) | Spe<br>(%) | AUC | Cut-off point<br>(≤) | Sens<br>(%) | Spe<br>(%) |
| ***HGS (kg)*** |  |  |  |  |  |  |  |  |  |
| /forearm circumference | (cm) | 0.74* | 1.33 | 86 | 59 | 0.70* | 1.04 | 84 | 58 |
| ***1RM<sub>knee extensors</sub> (kg)*** |  |  |  |  |  |  |  |  |  |
| /body mass | (kg) | 0.77* | 0.85 | 86 | 68 | 0.76† | 0.54 | 68 | 78 |
| /forearm circumference |  | 0.75* | 2.16 | 86 | 77 | 0.70* | 1.38 | 60 | 75 |
| /calf circumference | (cm) | 0.74* | 1.65 | 86 | 77 | 0.70* | 1.06 | 72 | 68 |
| /chest circumference |  | 0.76* | 0.64 | 86 | 73 | 0.71* | 0.4 | 72 | 65 |
| /waist circumference |  | 0.78* | 0.73 | 100 | 59 | 0.72* | 0.37 | 60 | 80 |
| /triceps skinfold |  | 0.81† | 4.22 | 86 | 68 | 0.70* | 1.40 | 60 | 75 |
| /bitrochanteric breadth | (mm) | 0.73* | 1.72 | 86 | 77 | 0.70* | 1.16 | 76 | 60 |
| /bimalleolar breadth |  | 0.73* | 8.77 | 86 | 77 | 0.70* | 5.77 | 76 | 65 |
| /elbow breadth |  | 0.73* | 9.36 | 86 | 73 | 0.70* | 6.57 | 76 | 63 |
| /SA | (m²) | 0.75* | 31.6 | 86 | 77 | 0.72* | 21.2 | 72 | 68 |
| /MAMC | (cm) | 0.77* | 2.42 | 86 | 73 | 0.72* | 1.54 | 64 | 75 |
| /CAMA | (cm²) | 0.71* | 2.03 | 100 | 41 | 0.73* | 0.90 | 52 | 93 |
| /FFM<sup>(LEAN et al., 1996)</sup> | (kg) | 0.76* | 1.11 | 86 | 77 | 0.72* | 1.00 | 72 | 68 |
| /FFM<sup>(BAUMGARTNER et al., 1991)</sup> | (kg) | 0.78* | 1.13 | 83 | 73 | 0.75† | 0.83 | 76 | 64 |
| /body mass<sup>0.44</sup> |  | 0.75* | 9.04 | 86 | 77 | 0.71* | 6.03 | 76 | 63 |
| /body mass<sup>0.67 (JARIC, 2003)</sup> | (kg) | 0.78* | 3.40 | 86 | 77 | 0.73† | 2.28 | 76 | 65 |
| /body mass<sup>0.96 (ABDALLA et al., 2020)</sup> |  | 0.77* | 1.00 | 86 | 68 | 0.75† | 0.45 | 44 | 100 |
| /body mass<sup>0.69 (ABDALLA et al., 2020)</sup> |  | 0.78* | 3.06 | 86 | 77 | 0.73* | 1.48 | 44 | 98 |
| /calf circumference<sup>1.10</sup> | (cm) | 0.75* | 1.14 | 86 | 77 | 0.71* | 0.70 | 68 | 73 |
| /bimalleolar breadth<sup>1.20</sup> | (mm) | 0.71* | 6.01 | 86 | 68 | 0.70* | 3.93 | 76 | 65 |
| /(body mass*height)<sup>0.48</sup> | (kg*m) | 0.75* | 5.83 | 86 | 77 | 0.72* | 4.06 | 76 | 65 |
| /SA<sup>0.93</sup> | (m²) | 0.75* | 33 | 86 | 77 | 0.72* | 22.7 | 76 | 65 |
| /FFM<sup>0.88 (LEAN et al., 1996)</sup> | (kg) | 0.75* | 1.77 | 86 | 77 | 0.71* | 1.53 | 76 | 65 |
| /FFM<sup>0.67 (BAUMGARTNER et al., 1991)</sup> |  | 0.76* | 3.94 | 83 | 77 | 0.72* | 2.91 | 76 | 64 |
| ***knee extensors PT<sup>60°/s</sup> (Nm)*** |  |  |  |  |  |  |  |  |  |
| /height | (m) | 0.93† | 54.1 | 86 | 95 | 0.74† | 44.1 | 64 | 75 |
| /knee height | (cm) | 0.94† | 1.83 | 100 | 86 | 0.75† | 1.44 | 68 | 73 |
| /SA | (m²) | 0.94† | 56.9 | 100 | 82 | 0.81† | 36.3 | 64 | 88 |
| /FFM<sup>(LEAN et al., 1996)</sup> | (kg) | 0.93† | 1.79 | 86 | 95 | 0.81† | 1.84 | 76 | 80 |
| /left leg LST |  | 0.82† | 0.015 | 100 | 55 | 0.76† | 0.012 | 68 | 80 |
| /right leg LST | (g) | 0.77* | 0.015 | 100 | 50 | 0.78† | 0.013 | 76 | 78 |
| /legs LST |  | 0.81* | 0.0063 | 71 | 82 | 0.77† | 0.0065 | 76 | 78 |
| /ASM |  | 0.81* | 4.66 | 71 | 86 | 0.77† | 5.01 | 76 | 78 |
| /FFM<sup>(BAUMGARTNER et al., 1991)</sup> | (kg) | 0.93† | 1.63 | 83 | 95 | 0.85† | 1.60 | 88 | 77 |
| /FFM<sup>(DXA)</sup> |  | 0.84† | 2.08 | 100 | 59 | 0.79† | 1.89 | 76 | 75 |
| /body mass<sup>0.67 (DAVIES; DALSKY, 1997)</sup> |  | 0.93† | 5.06 | 86 | 95 | 0.82† | 3.71 | 68 | 88 |
| /body mass<sup>0.72 (DAVIES; DALSKY, 1997)</sup> | (kg) | 0.94† | 4.1 | 86 | 95 | 0.82† | 3.14 | 72 | 85 |
| /body mass<sup>0.74 (DAVIES; DALSKY, 1997)</sup> |  | 0.93† | 3.77 | 86 | 95 | 0.82† | 2.87 | 72 | 85 |
| /body mass<sup>0.67 (JARIC, 2003)</sup> |  | 0.93† | 5.06 | 86 | 95 | 0.82† | 3.71 | 68 | 88 |
| /height<sup>3.27</sup> | (m) | 0.86† | 19.2 | 100 | 77 | 0.74† | 17.4 | 72 | 65 |
| /knee height<sup>1.82</sup> | (cm) | 0.90† | 0.068 | 100 | 86 | 0.74† | 0.053 | 64 | 75 |
| /half arm span<sup>1.62</sup> |  | 0.88† | 0.076 | 100 | 73 | 0.75† | 0.066 | 84 | 58 |
| /biacromial breadth<sup>2.15</sup> | (mm) | 0.82† | 0.038 | 86 | 68 | 0.77† | 0.03 | 76 | 73 |
| /bimalleolar breadth<sup>1.54</sup> |  | 0.81* | 5.04 | 86 | 73 | 0.80† | 4.64 | 92 | 60 |
| /(body mass*height)<sup>0.43</sup> | (kg*m) | 0.94† | 13 | 100 | 82 | 0.80† | 10.5 | 84 | 65 |
| /SA<sup>0.83</sup> | (m²) | 0.94† | 53.8 | 86 | 95 | 0.80† | 50 | 84 | 65 |
| /left leg LST<sup>0.43</sup> |  | 0.92† | 2.26 | 100 | 77 | 0.77† | 1.83 | 76 | 70 |
| /right leg LST<sup>0.48</sup> | (g) | 0.90† | 1.39 | 100 | 73 | 0.78† | 1.16 | 80 | 70 |
| /legs LST<sup>0.47</sup> |  | 0.92† | 1.07 | 100 | 73 | 0.77† | 0.87 | 76 | 70 |

\* $p < 0.05$ and † $p < 0.001$ (statistically significant AUC)
Note: AUC=area under the curve; $p$ =significance; Sens=sensitivity; Spe=specificity; SA= human body surface area; MAMC=mid-arm muscle circumference; CAMA=corrected arm muscle area; FFM=Fat-free mass; LST=lean soft tissue; ASM=appendicular skeletal muscle mass; DXA=Dual-energy X-ray absorptiometry; 6MWT=six-minute walk test.

A comparison of the most accurate ROC curves is presented in Figure 2 to support the decision for the best cut-off point between non-normalized, ratio standard and allometric scaling of HGS and lower limbs strength (1RM_knee extensors_ and _knee extension_PT^60°/s^) for each sex.

**Figure 2.**
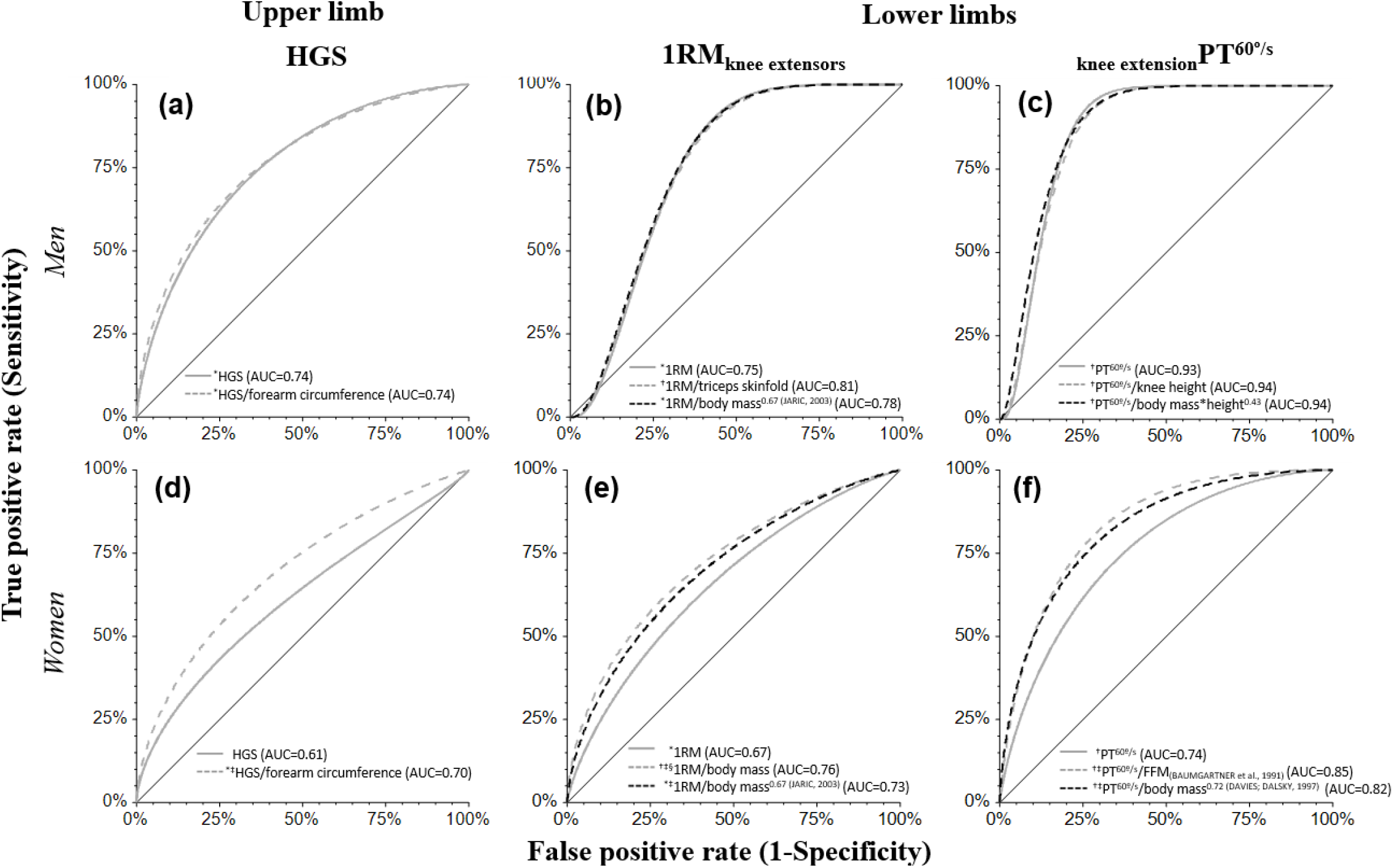
Accuracy comparison between non-normalized, ratio standard and allometric scaling of muscle weakness cut-off points of HGS and lower limbs strength (1RM_knee extensors_ and _knee extension_PT^60°/s^) in older men (letters a, b, c) and older women (letters d, e, f). ^*^p<0.05 and ^†^p<0.001 (statistically significant AUC). ^‡^p<0.05 (greater than the AUC of non-normalized muscle strength). ^§^ p<0.05 (greater than the AUC of the allometric scaling). Dependent variable (primary outcome): functional limitation (6MWT<400 m). *Note*: HGS=handgrip strength; 1RM_knee extensors_=one maximum repetition measurement for knee extensors; _knee extension_PT^60°/s^=isokinetic knee extension peak torque at 60°/s; 6MWT=six-minute walk tes

For men, there were no differences in accuracy (AUC) to identify functional limitation between absolute muscle strength, normalized by ratio standard or by allometric scaling (p>0.05; Figure 2 a, b, c). However, the absolute muscle strengths (HGS, 1RM_knee extensors_ and _knee extension_PT^60°/s^) previously indicated great dependence (r>0.30) on body size (SUPPLEMENT C), suggesting the need for normalization to avoid errors in the classification of weakness. The normalized muscle strength increased the AUC and made it possible to classify muscle weakness of older adults with extreme body sizes, independently.

For women, only after normalizing muscle strength the AUC values perform acceptable to identify functional limitation (AUC>0.70; Figure 2 d, e). The exception was _knee extension_PT^60°/s^, when the absolute values already had adequate accuracy (AUC>0.70), although without the desirable independence of body size. All the normalizations increased (^‡^) the AUC (p<0.001).

## DISCUSSION

Cut-off points based on upper and lower limbs muscle strength were proposed to identify muscle weakness in older adults of both sexes. The non-normalized cut-off points for HGS and lower limbs strength were significantly associated with body size, which involves biases to assess older adults with extreme body size (e.g., heavy or short). After normalizing HGS and lower limbs strength by the ratio standard or by the allometry, the association with body size was no longer relevant. In addition, for women, the accuracy to predict mobility limitation/muscle weakness from normalized muscle strength cut-off points become acceptable when compared to non-normalized strategy. In men, muscle strength normalization did not increase accuracy. However, all normalized models of both sexes avoided biases in the assessment of muscle weakness/mobility limitation, to isolate the natural interdependence between muscle strength and body size [21].

To the best of our knowledge, this is the first study to propose muscle weakness cut-off points for the HGS and _knee extension_PT^60°/s^ allometrically adjusted in older adults. In a previous study, 1RM_knee extensors_ was allometrically adjusted for body mass [23], but not according to all other potential body size variables. Indeed, we expanded the number of variables that can be used to normalize 1RM_knee extensors_ with allometry (n=8) in order to augment model’s accuracy for identifying muscle weakness regardless of extreme body sizes. Other studies proposed muscle weakness cut-off points with HGS normalized by ratio standard (body mass or BMI) [18, 54, 55] or stratified by BMI quartiles [7]. There are also muscle weakness cut-off points for _knee extension_PT^60°/s^ normalized by body mass [19]. However, these studies did not compare the accuracy of normalized with non-normalized muscle strength to identify muscle weakness. Furthermore, they did not explore other body-size variables to normalize muscle strength.

Previous studies have proposed allometric exponents to normalize muscle strength, including HGS [20-22], 1RM_knee extensors_ [23], _knee extension_PT^60°/s^ [25, 34], and they are comparable with the ones found in the present study. Curvilinear (allometric) relationship variables is confirmed when allometric coefficient (^b^) is between 0.00 and 0.99 [56], while the linear relationship is characterized when the exponent is ≥1.00 [56]. In the literature, body mass generally presents an allometric relationship with muscle strength independently of the test (HGS, 1RM_knee extensors_ or _knee extension_PT^60°/s^; Table 1), confirming our findings (SUPPLEMENT B), when ^b^ exponents were 0.22 (HGS), 0.44 (1RM_knee extensors_) and 0.37 (_knee extension_PT^60°/s^). Contrarily, height tends to have a linear relationship (^b^≥1.00) with muscle strength [21], what was also confirmed by our proposed allometric exponents (SUPPLEMENT B), that were between 1.87 and 3.27.

Some strengths of our study are noteworthy. We proposed muscle weakness cut-off points for isokinetic dynamometer, considered as a “ gold standard” resource to assess lower limbs strength. The estimated 1RM_knee extensors_ obtained with submaximal repetition protocol and the HGS are valid for older adults, even for those with muscle weakness [48, 57]. An extensive number of body-size variables (n=49) were tested in our study, expanding the possibilities to promote the normalization of performance in muscle strength tests. Furthermore, regardless of the chosen muscle strength test to evaluate weakness, our findings can be applied with sufficient accuracy (AUC>0.70) both for scientific research (_knee extension_PT^60°/s^) and population-based monitoring (HGS and 1RM_knee extensors_). Nevertheless, this study is not without limitations. The individual muscle strength decline along aging may have been underestimated with the cross-sectional design. Another limitation is the small and local sample size of our study, requiring caution to extrapolate these findings inferentially to other populations.

We found greater accuracy (AUC) for normalized lower limbs strength (isokinetic dynamometer and leg extension machine) than manual dynamometer (normalized upper extremity strength), usually adopted to predict mobility limitations/muscle weakness [8]. However, the isokinetic dynamometer is expensive and generally more available in terms of research. Even though, our normalized models are also applicable in clinical practice from manual dynamometers (widely available in geriatric environments) and leg extension machines (available in most fitness centers, adequate environment for intervention against aged-related muscle weakness) [30]. The assessment of HGS and 1RM_knee extensors_ and proper classification of muscle weakness amongst older adults should be frequent in clinical practice to better target health expending, avoiding unnecessary expenditures. Future research should observe if proposed allometric exponents can be utilized to normalize muscle strength for different older adults’ population, with other ethnicity/race characteristics.

As an applied example to avoid false positive diagnosis for muscle weakness, we hypothesize one older man with extreme lower values of body mass (42 kg), 1.57 m of height, who performed _knee extension_PT^60°/s^ of 85.2 Nm. If we consider our absolute cut-off point (≤85.4 Nm), this older man has muscle weakness confirmed. However, when considered the normalized _knee extension_PT^60°/s^/([body mass*height]^0.43^), the adjusted value (14.1 Nm/kg*m) is above of the cut-off point (13.0 Nm/kg*m; Table 3). Normalization would also avoid false negative cases, for large body size of older adults. The mistaken framing of false positive weakness cases could greatly impact the financial resources in the health and older people care systems. Especially in low- and middle-income countries, where these resources are scarcer.

## CONCLUSION

Upper and lower limbs muscle weakness cut-off points standardized according to body size were proposed for older adults of both sexes. The normalization has increased accuracy for identify women with muscle weakness; but not in men, whose absolute muscle strength values have an acceptable accuracy. However, normalization made muscle strength independent of body size, confirming our hypothesis and preventing bias in the evaluation of older adults with extreme body size (e.g., very low or very heavy). Forty-nine valid models were proposed for older adults of both sexes, with different possibilities of body’s normalization of muscle strength, which broadens the interpretation of muscle strength with less risk of attributing a false-positive diagnosis to muscle weakness.

## Data Availability

The data that support the findings of this study are available from the corresponding author, [PPA], upon reasonable request.

## Acknowledgements

Nothing to declare.

## SUPPLEMENTS

**SUPPLEMENT A.** Body size variables (n=49) to normalize muscle strength.

| **Anthropometry** |  |  |
| --- | --- | --- |
| Body mass (kg) |  |  |
| Height (m) |  |  |
| *Circumferences (cm)* | *Skinfold site (mm)* | *Bone breadths (mm)* |
| Arm | Subscapular | Biacromial |
| Forearm | Triceps | Biiliac |
| Midthigh | Biceps | Bitrochanteric |
| Calf | Midaxillary | Ankle (bimalleolar) |
| Chest | Pectoral (chest) | Elbow |
| Waist | Suprailiac | Wrist |
| Abdomen | Abdominal (vertical) | Knee |
| Buttocks (hip) | Thigh (midline) | Chest |
|  | Medial calf |  |
| *Segment lengths (cm)* |  |  |
| Knee height |  |  |
| Half arm span |  |  |
| **Body indexes** |  |  |
| *Derived from anthropometry* | *Derived from body composition* |  |
| BMI (kg/m<sup>2</sup>) | Left arm LST (g) |  |
| Body mass*height (kg*m) | Right arm LST (g) |  |
| SA (m<sup>2</sup>) | Left leg LST (g) |  |
| MAMC (cm) | Right leg LST (g) |  |
| CAMA (cm<sup>2</sup>) | Arms LST (g) |  |
| AFA (cm<sup>2</sup>) | Legs LST (g) |  |
| FFM<sub>(LEAN et al., 1996)</sub> (kg) | ASM (kg) |  |
| Fat mass<sub>(LEAN et al., 1996)</sub> (kg) | ASM/height<sup>2</sup> (kg/m<sup>2</sup>) |  |
|  | FFM<sub>(BAUMGARTNER et al., 1991)</sub> (kg) |  |
|  | FFM<sub>(DXA)</sub> (kg) |  |
|  | Fat mass<sub>(BAUMGARTNER et al., 1991)</sub> (kg) |  |
|  | Fat mass<sub>(DXA)</sub> (kg) |  |

*Note:* BMI=body mass index; SA=surface area of human body; MAMC=mid-arm muscle circumference; CAMA=corrected arm muscle area; AFA=arm fat area; FFM=fat-free mass; LST=lean soft tissue; ASM=appendicular skeletal muscle mass.
Instruments utilized for measures: body mass (Filizola® digital scale, model Personal, Brazil), height (Sanny® wall-mounted aluminum stadiometer, Professional model ES2020, Brazil), circumferences (Sanny® inelastic and inextensible measuring tape, Brazil), skinfolds (Lange scientific skinfold caliper, Cambridge Scientific Instruments, Cambridge, Maryland), bone breadths (Sanny® anthropometer and small sliding caliper, Brazil), and segment lengths (Sanny® segmometer, Brazil), LST components, ASM and FFM<sub>(DXA)</sub> (Dual Energy X-ray Absorptiometry-DXA, Hologic®, model QDR4500W, software version 11.2, Bedford, MA), FFM<sub>(BAUMGARTNER et al., 1991)</sub> (Bioimpedance Imp DF50 Body Composition Analysis, ImpediMed®, Brisbane, Queensland, Australia).

**SUPPLEMENT B.**
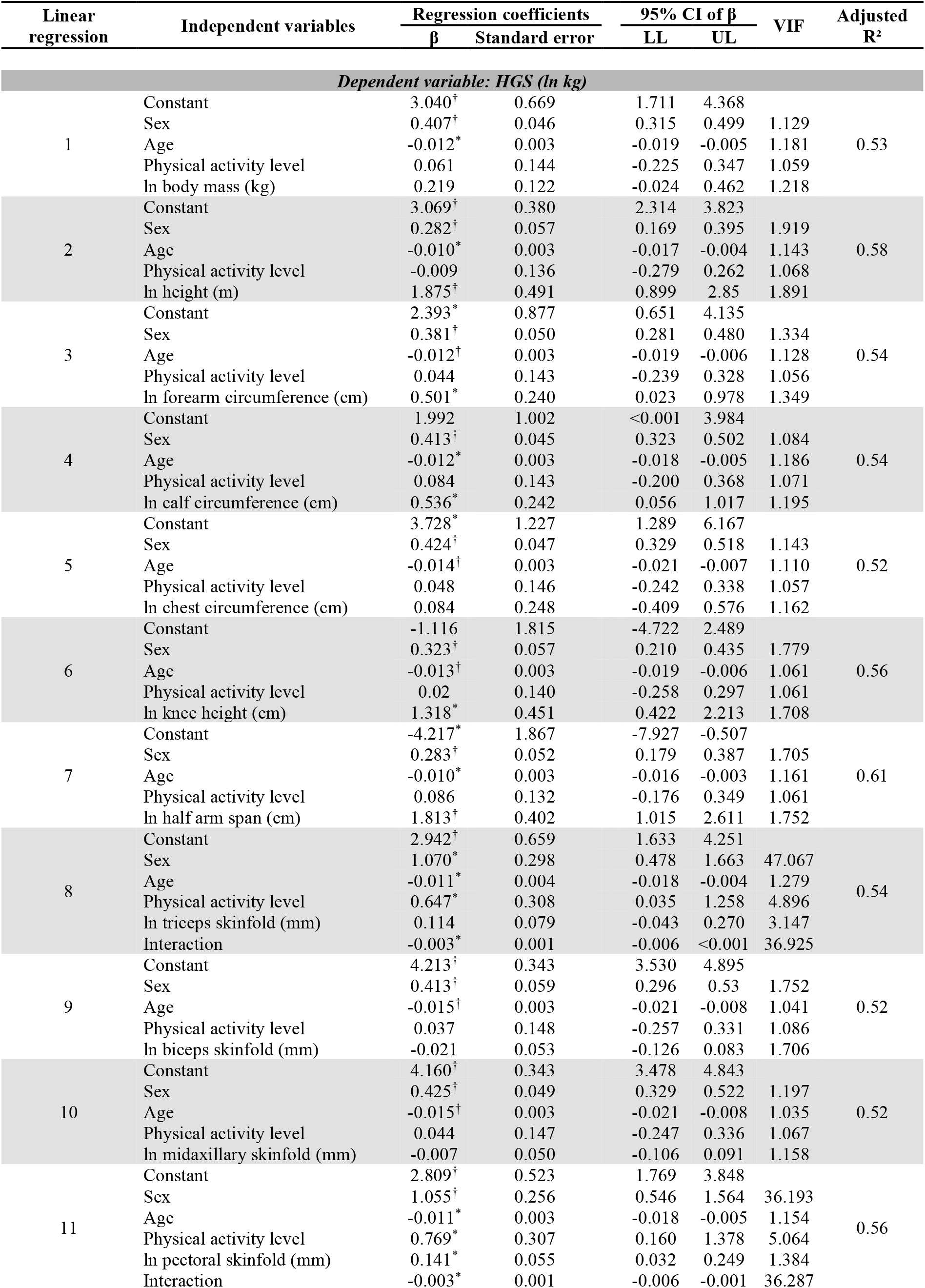
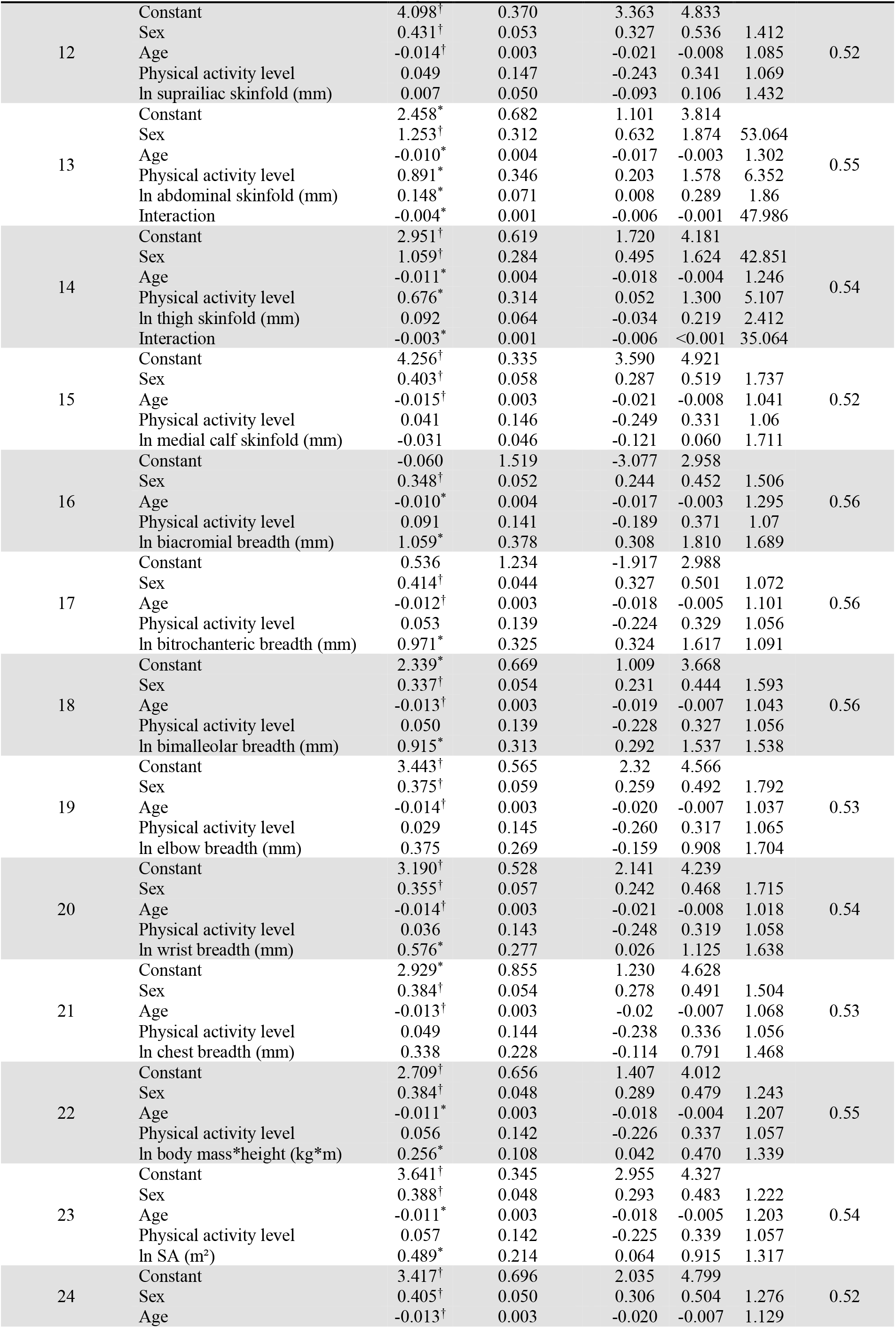
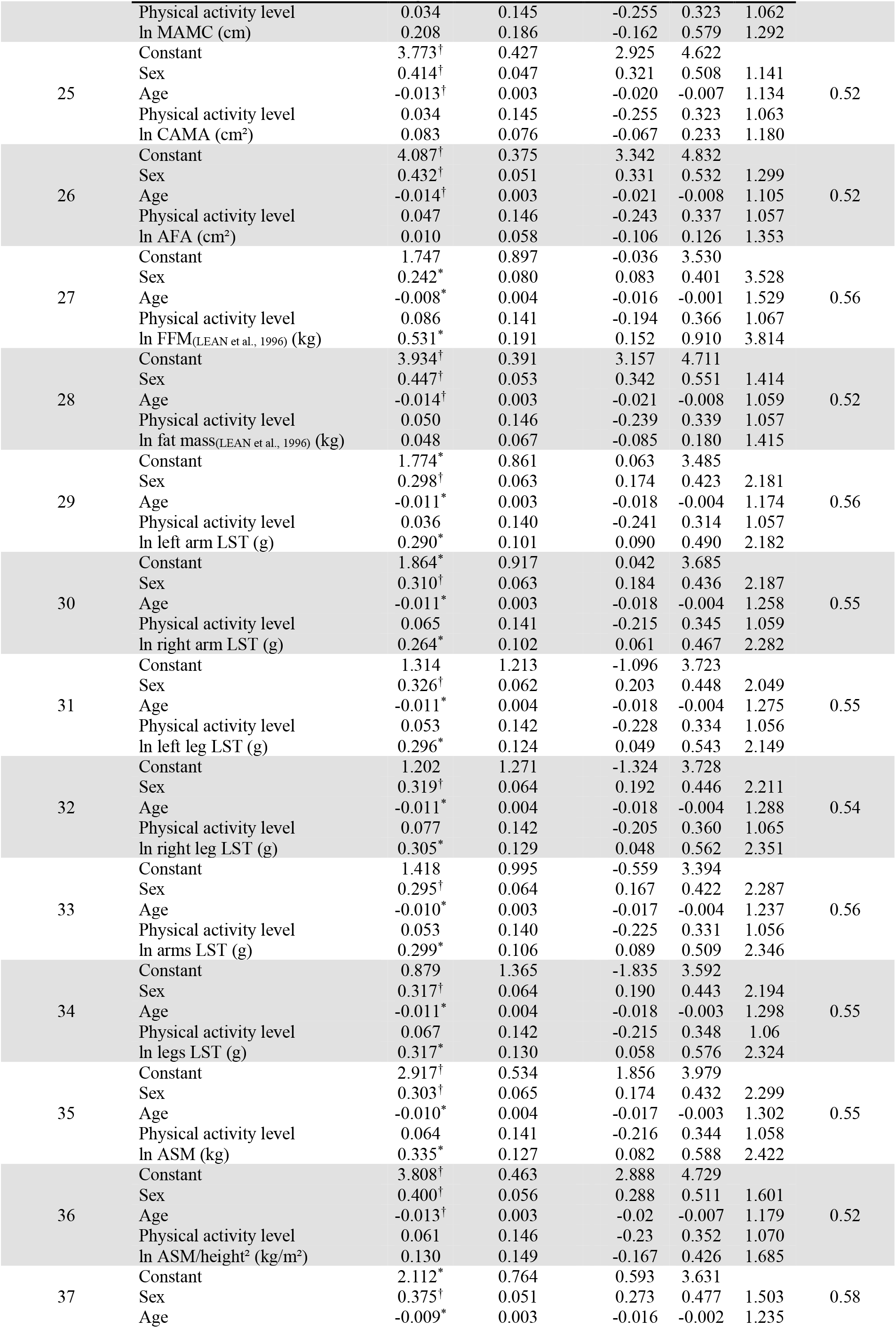
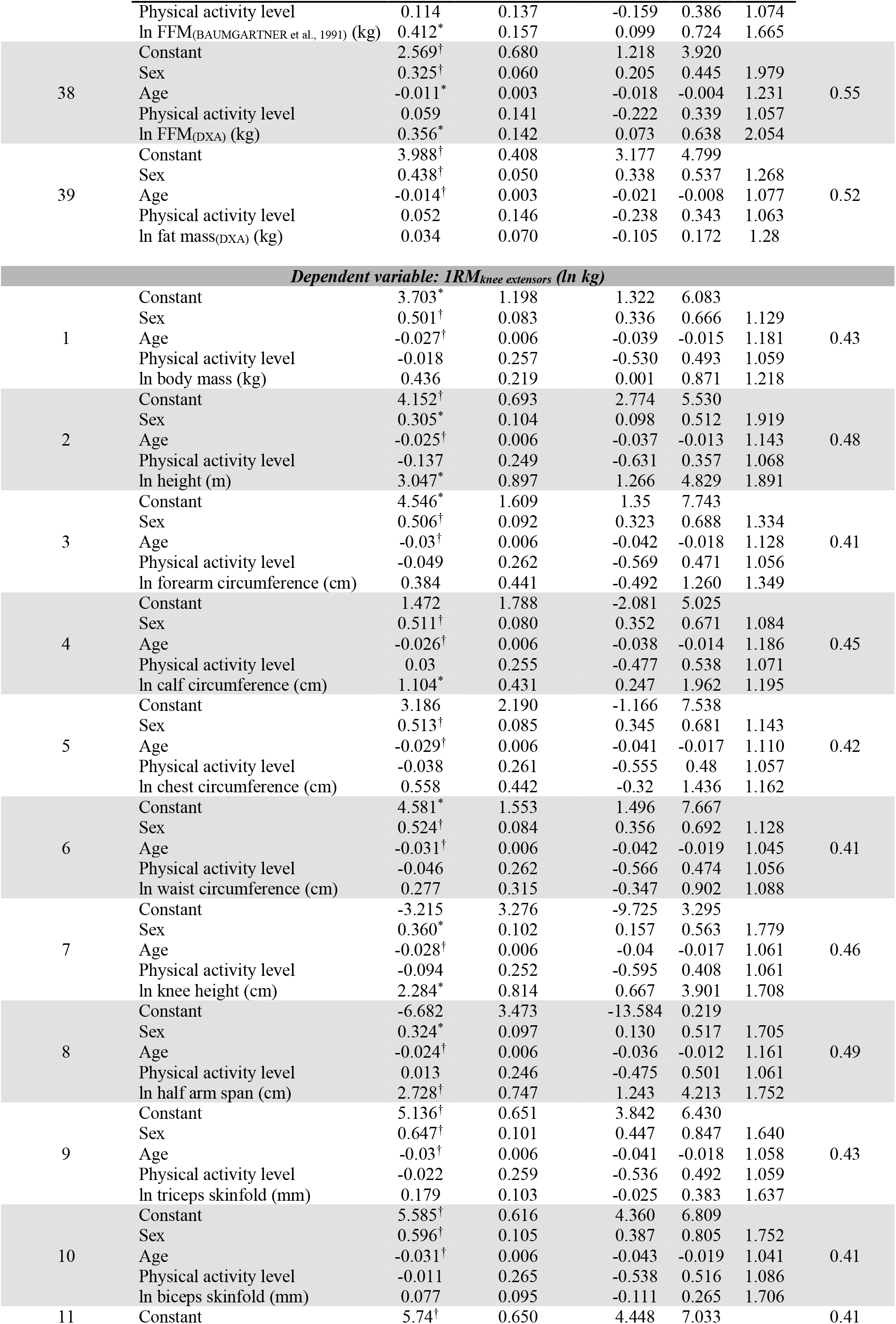
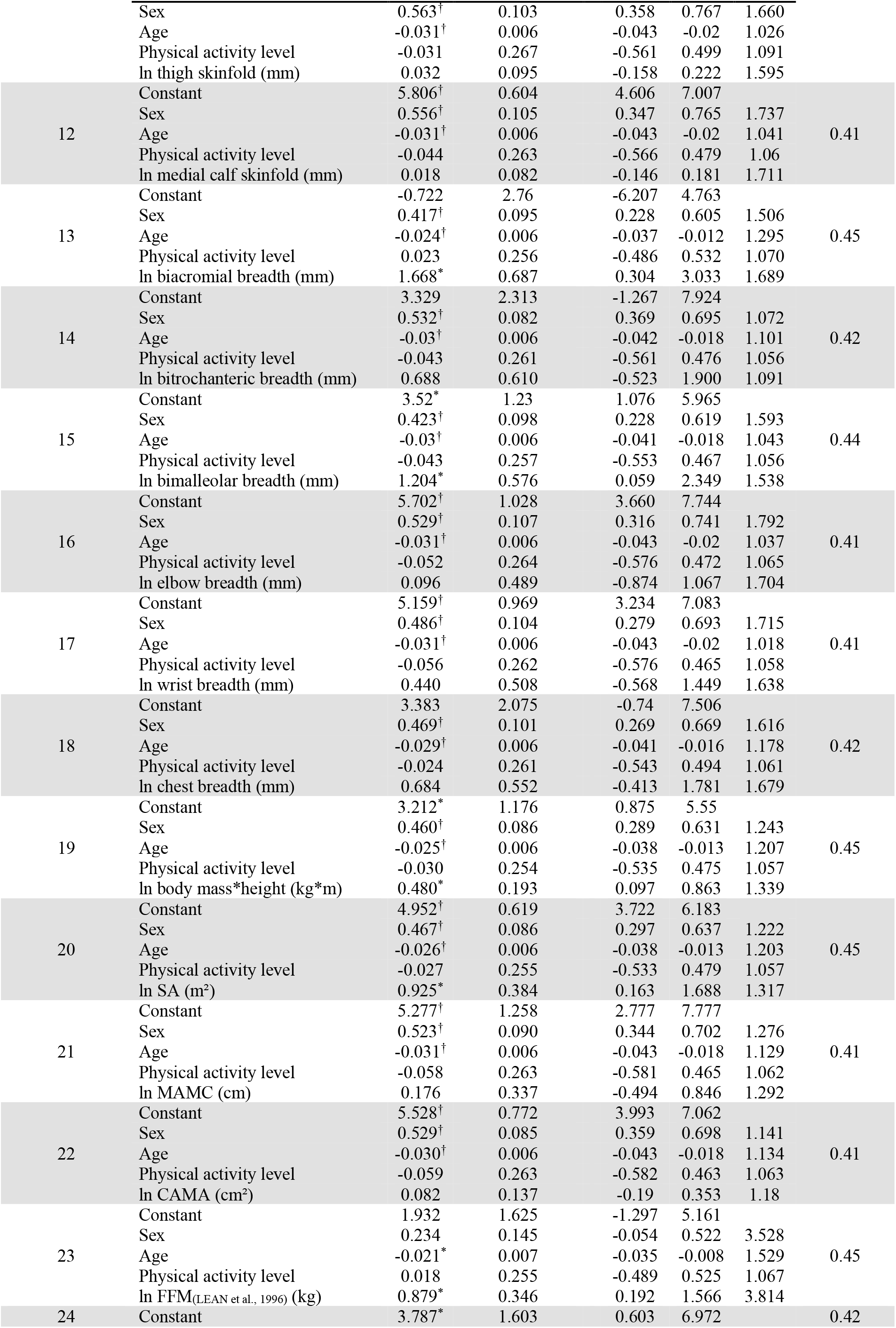
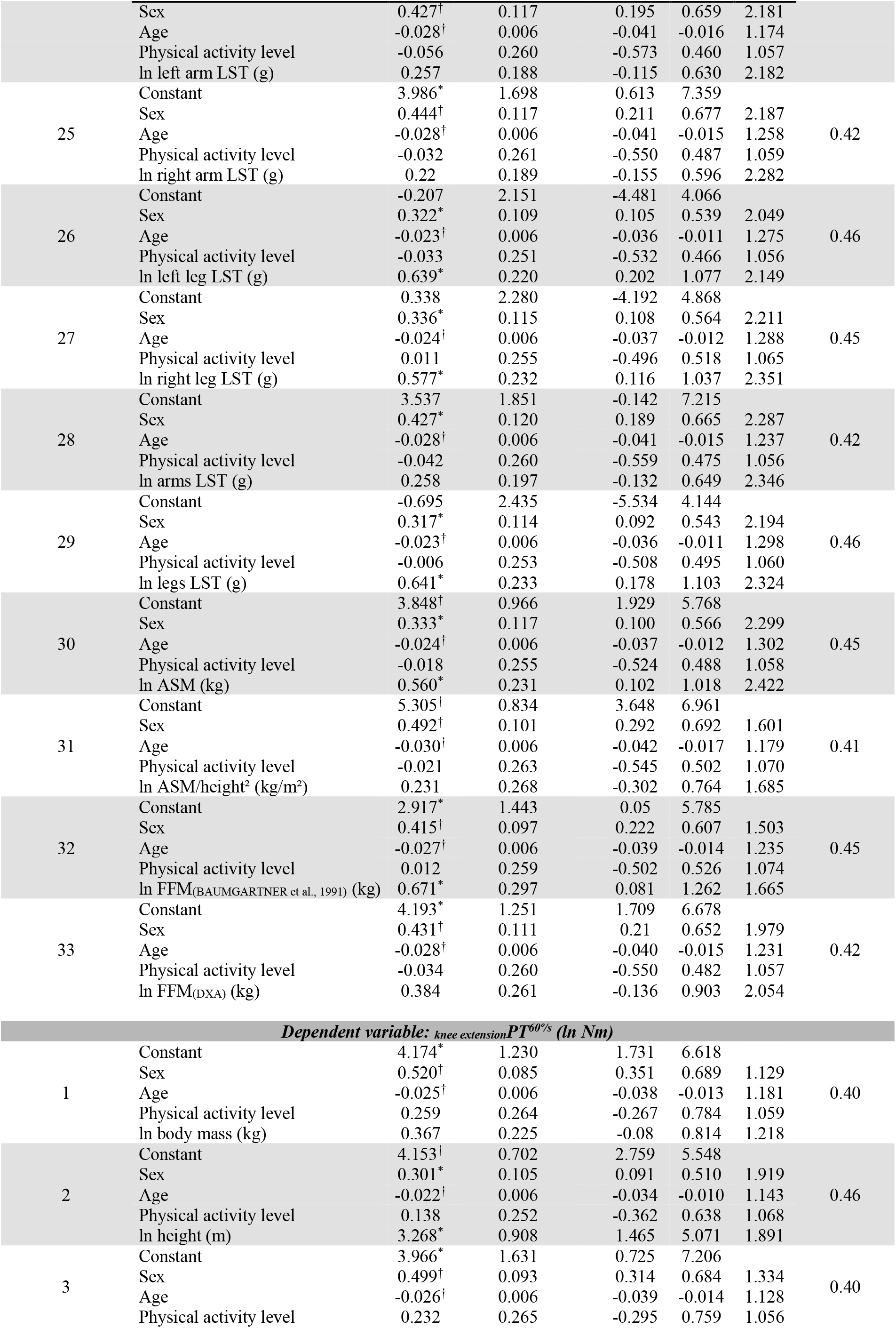
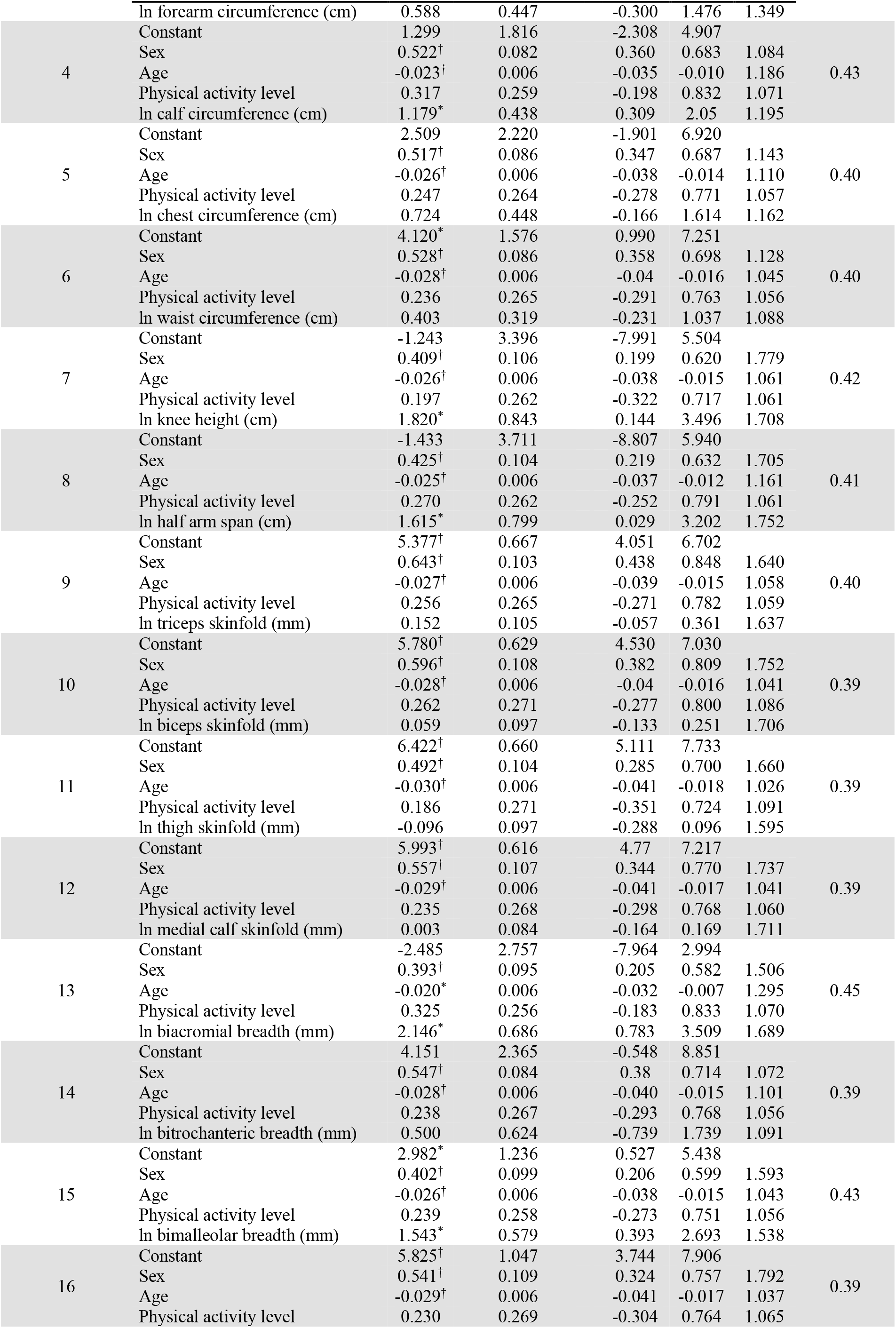
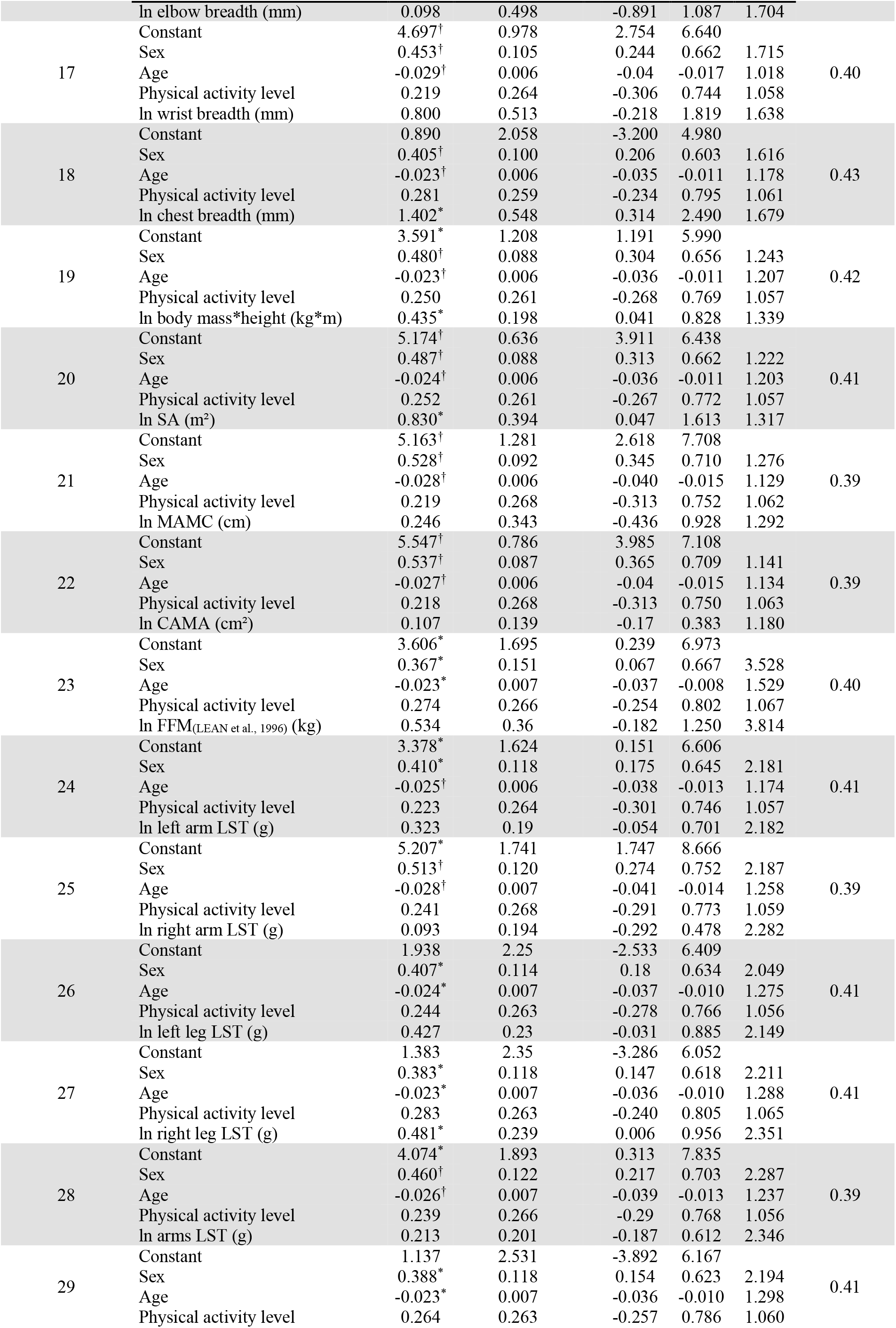
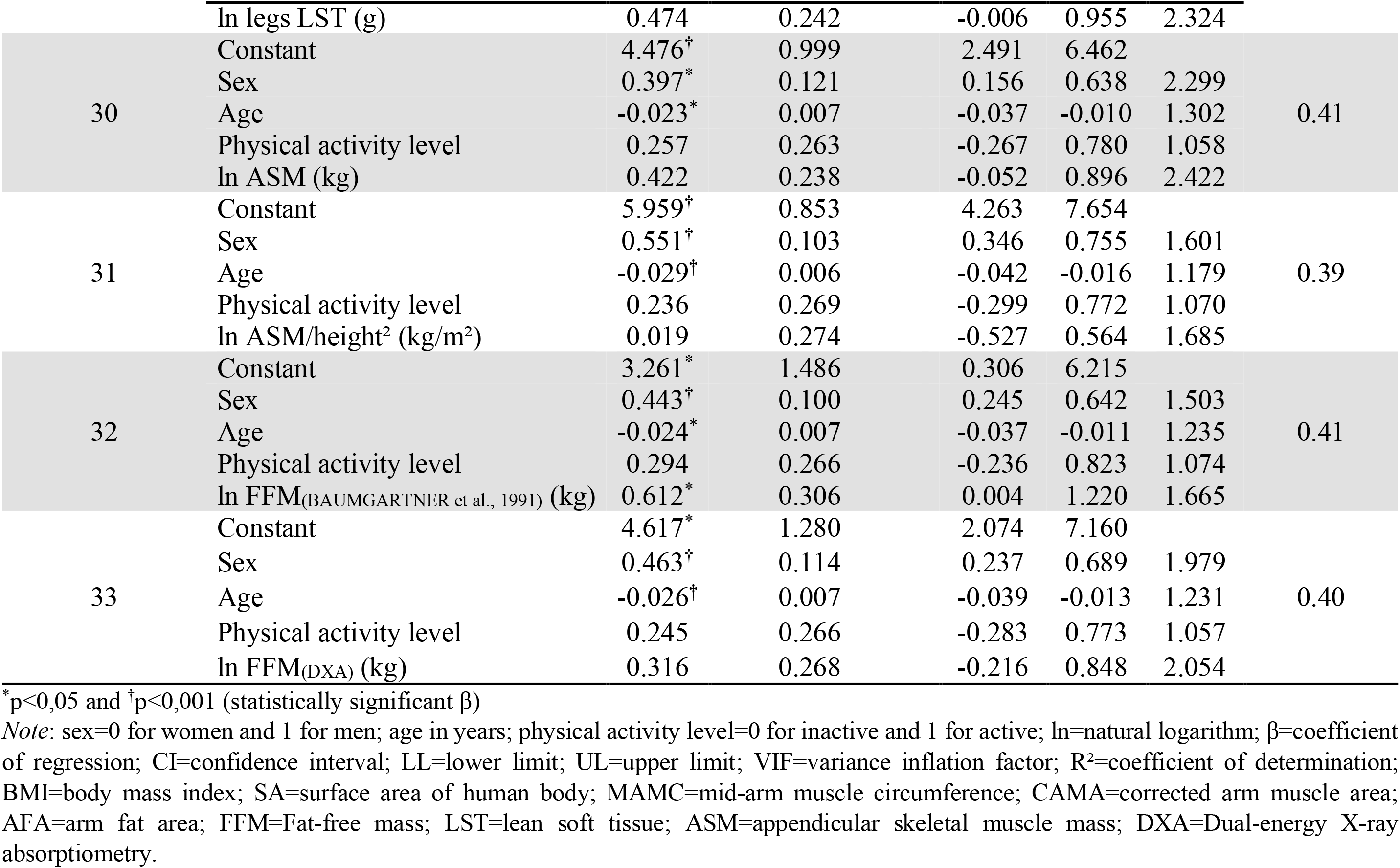
Linear regressions to obtain allometric exponents for handgrip strength (HGS), one maximum repetition measurement for knee extensors (1RM_knee extensors_) and isokinetic knee extension peak torque at 60°/s (_knee extension_PT^60°/s^) in older men and women (n=94).

**SUPPLEMENT C.**
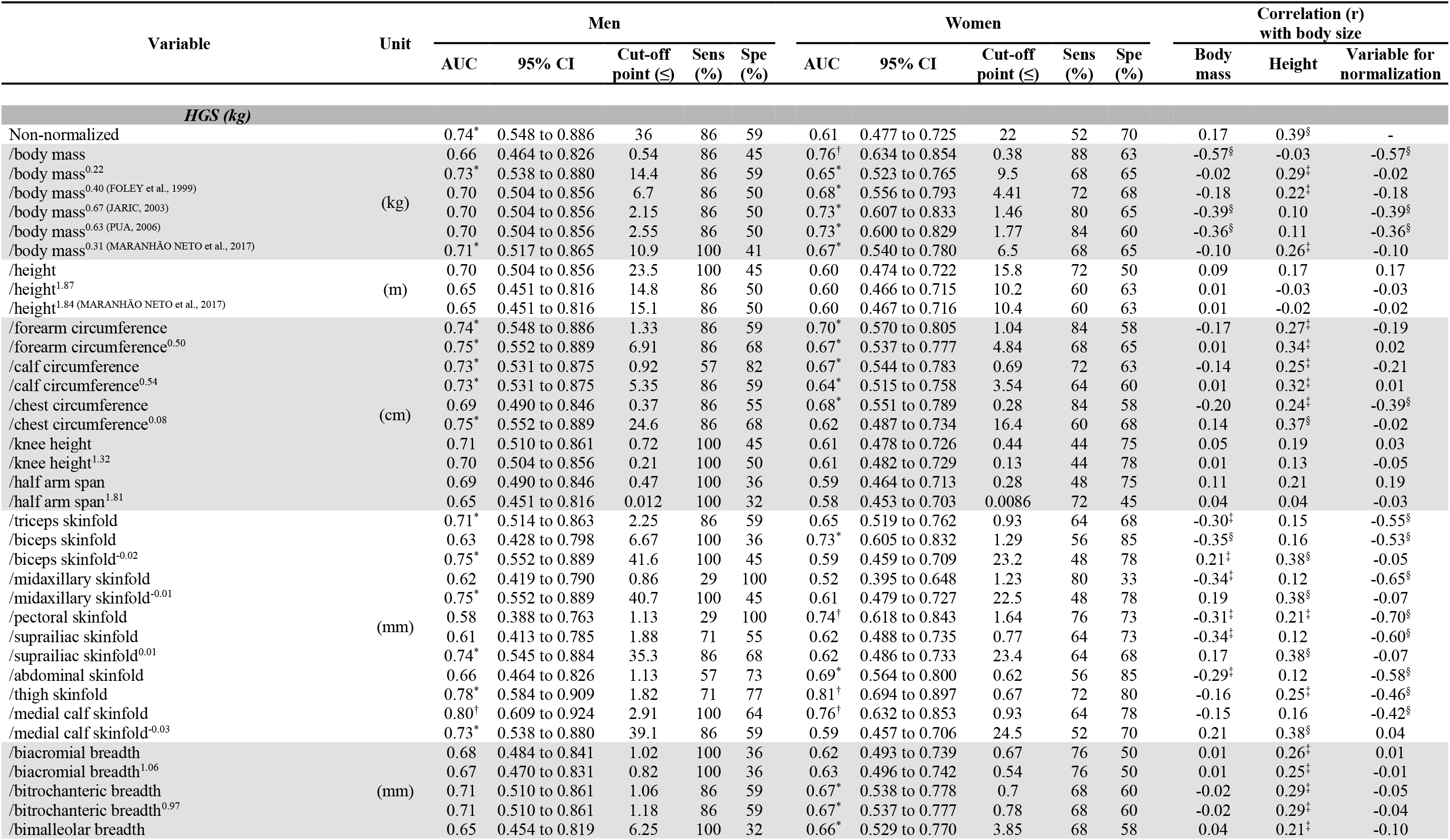
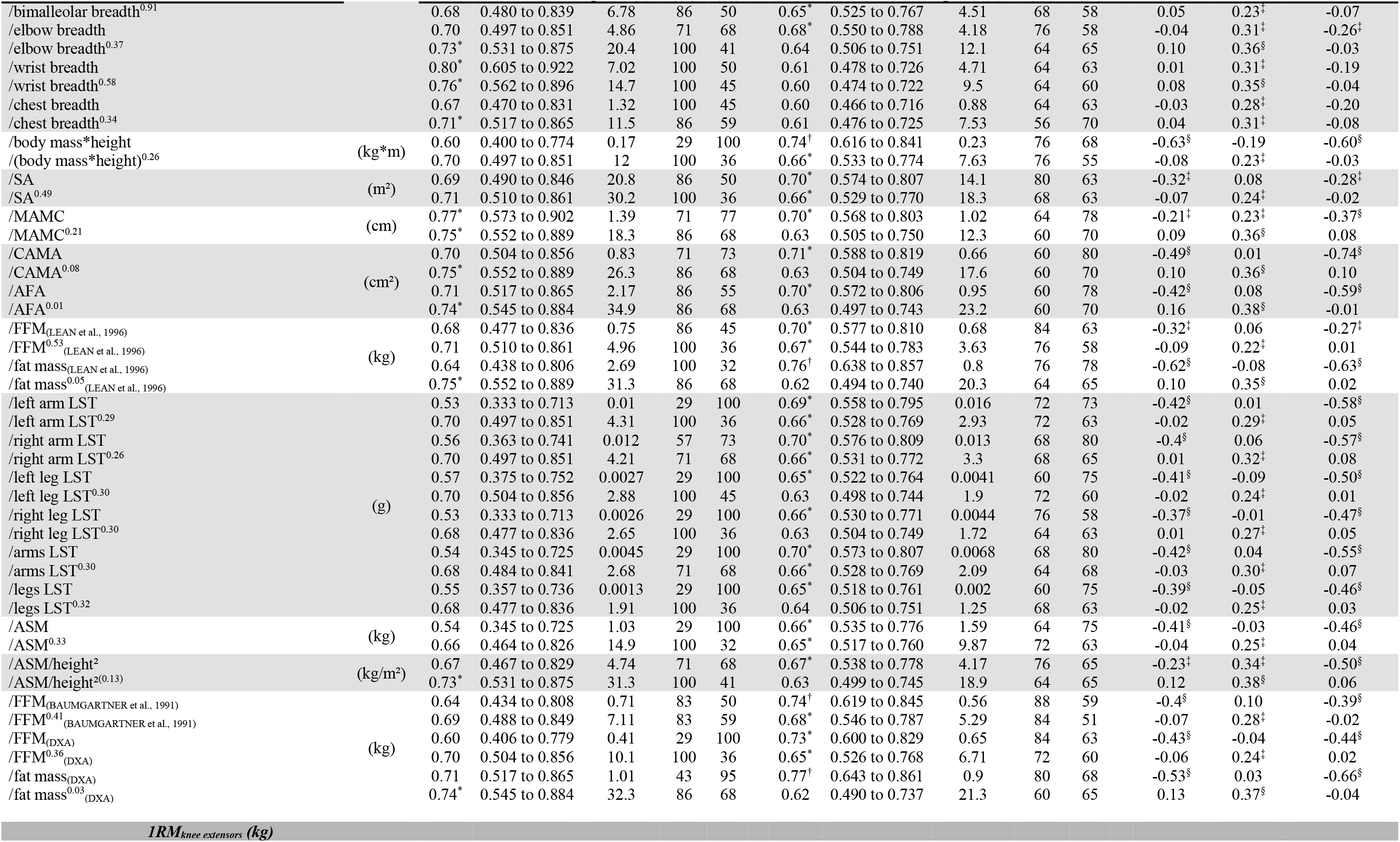
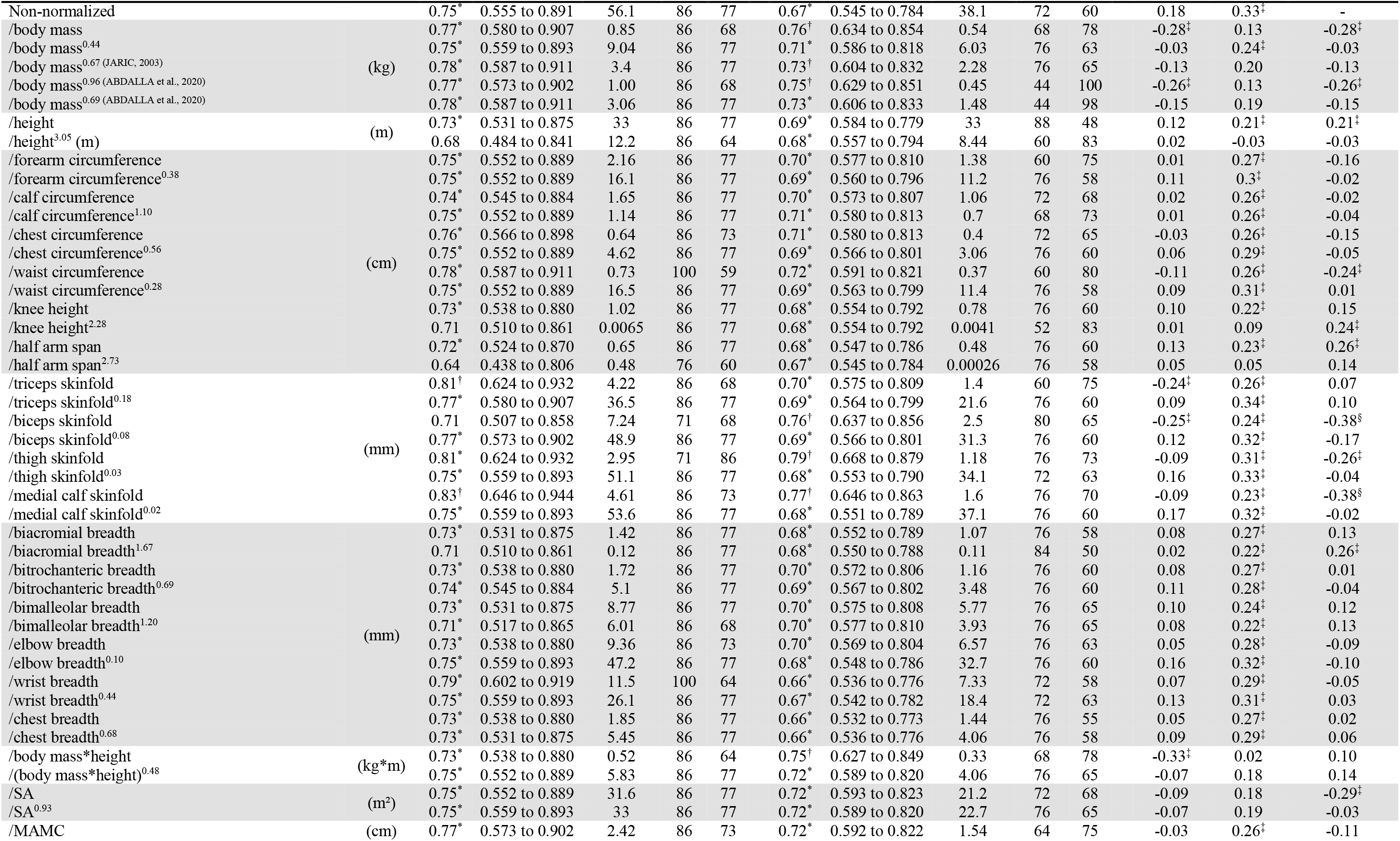
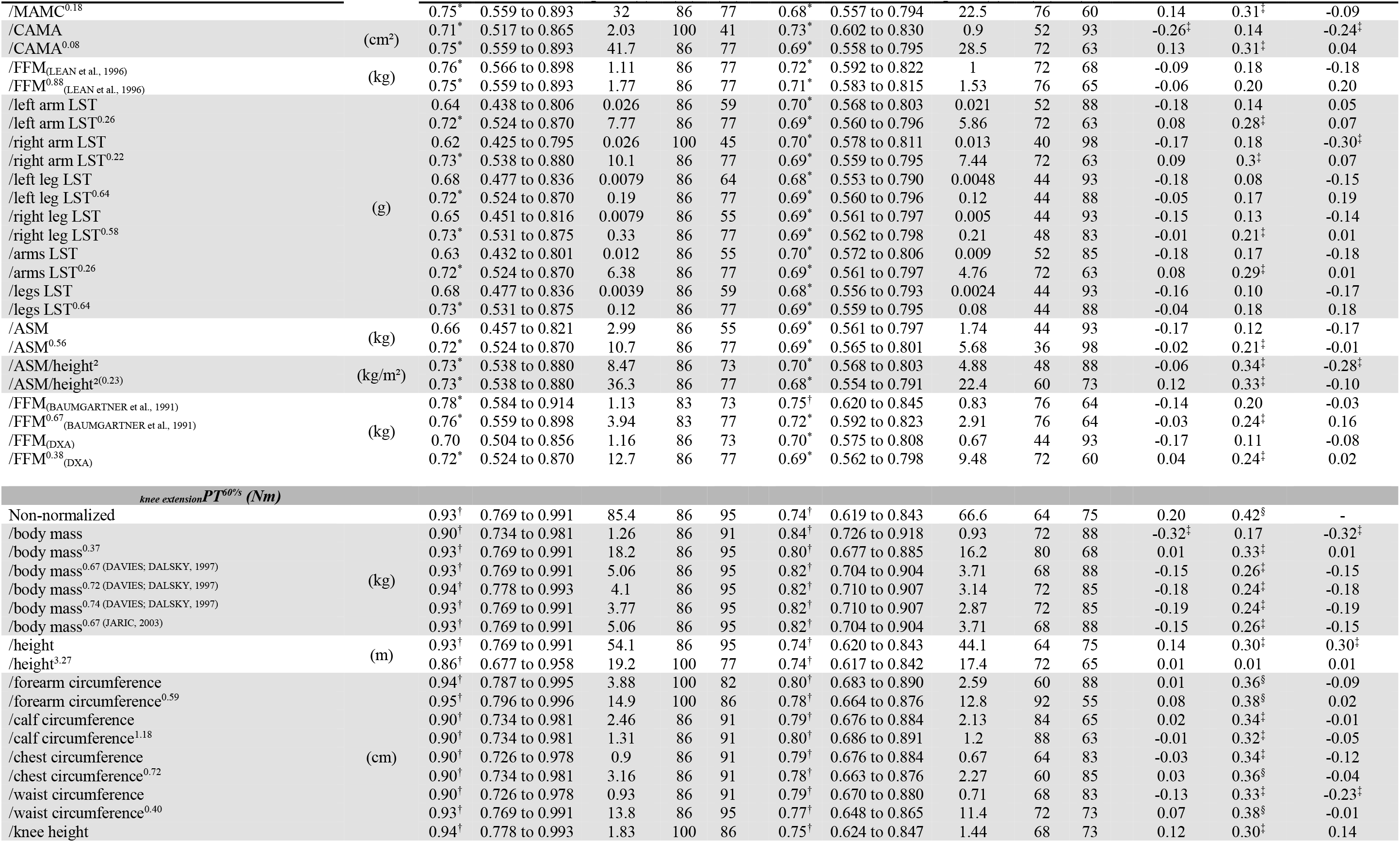
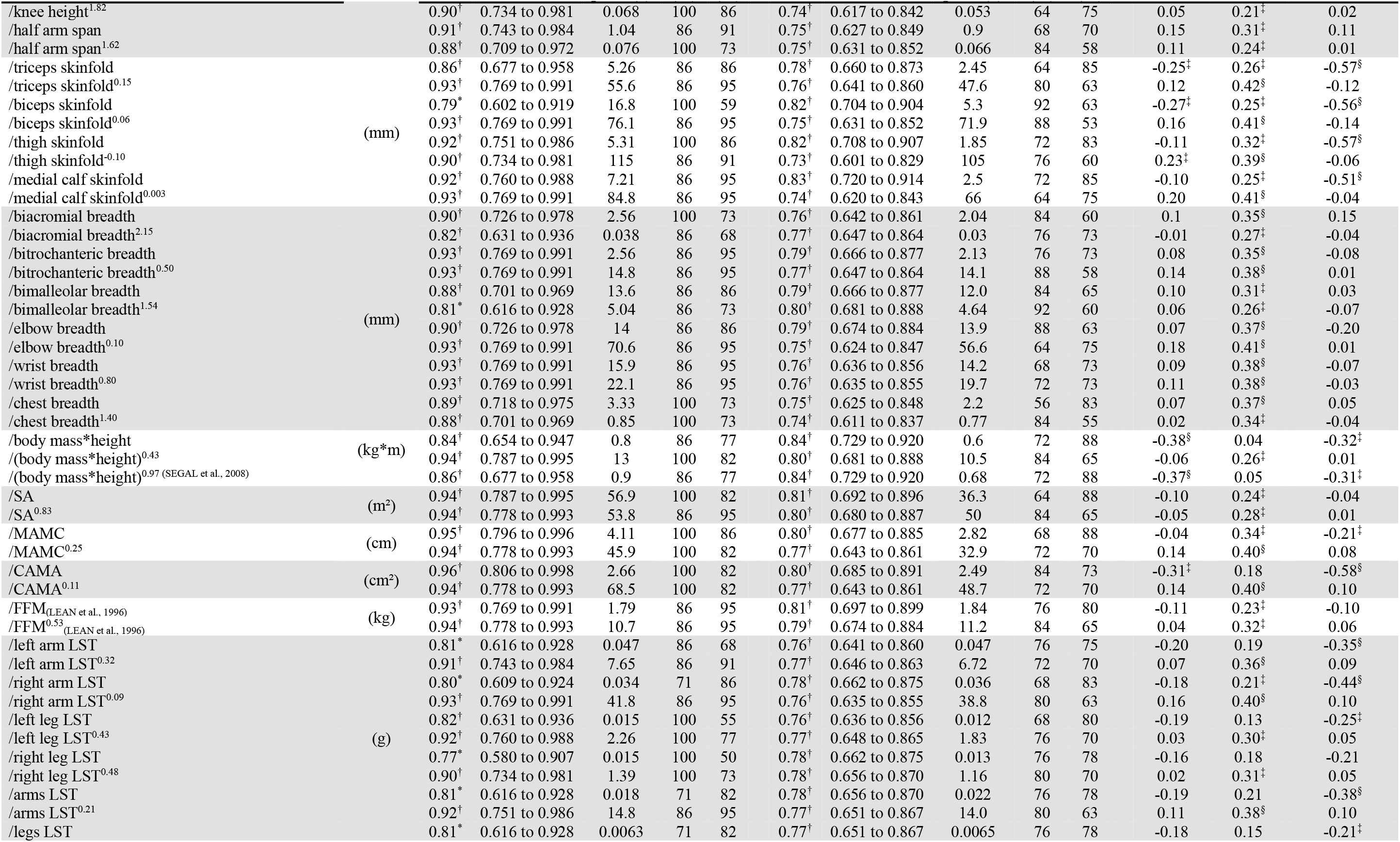
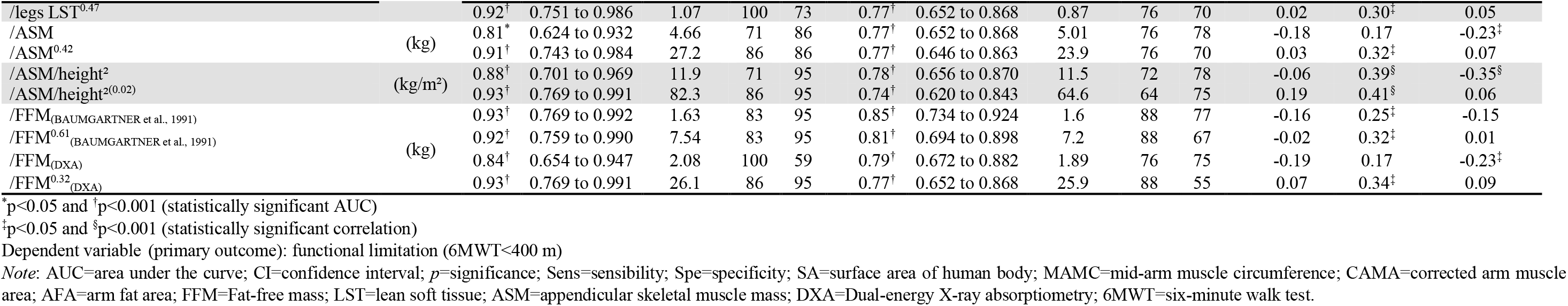
Cut-off points to identify muscle weakness in older adults of the handgrip strength (HGS), one maximum repetition measurement for knee extensors (1RM_knee extensors_) and isokinetic knee extension peak torque at 60°/s (_knee extension_PT^60°/s^) (non-normalized, ratio standard and allometric scaling), and the correlation of muscle strength with body size.

## REFERENCES

1. Bohannon RW. Hand-grip dynamometry predicts future outcomes in aging adults. Journal of geriatric physical therapy (2001). 2008;31:3-10. doi:10.1519/00139143-200831010-00002

2. Teng Z, Zhu Y, Yu X, Liu J, Long Q, Zeng Y, et al. An analysis and systematic review of sarcopenia increasing osteopenia risk. PloS one. 2021;16:e0250437. doi:10.1371/journal.pone.0250437

3. Bohannon RW. Grip Strength: An Indispensable Biomarker For Older Adults. Clinical interventions in aging. 2019;14:1681-91. doi:10.2147/cia.s194543

4. Santanasto AJ, Miljkovic I, Cvejkus RK, Wheeler VW, Zmuda JM. Sarcopenia Characteristics Are Associated with Incident Mobility Limitations in African Caribbean Men: The Tobago Longitudinal Study of Aging. J Gerontol A Biol Sci Med Sci. 2020;75:1346-52. doi:10.1093/gerona/glz233

5. Landi F, Liperoti R, Russo A, Capoluongo E, Barillaro C, Pahor M, et al. Disability, more than multimorbidity, was predictive of mortality among older persons aged 80 years and older. Journal of clinical epidemiology. 2010;63:752-9. doi:10.1016/j.jclinepi.2009.09.007

6. Clark BC, Manini TM. Sarcopenia ≠ Dynapenia. The Journals of Gerontology: Series A. 2008;63:829-34. doi:10.1093/gerona/63.8.829

7. Fried LP, Tangen CM, Walston J, Newman AB, Hirsch C, Gottdiener J, et al. Frailty in older adults evidence for a phenotype. J Gerontol A Biol Sci Med Sci. 2001;56:M146–M57.

8. Cruz-Jentoft AJ, Bahat G, Bauer J, Boirie Y, Bruyère O, Cederholm T, et al. Sarcopenia: revised European consensus on definition and diagnosis. Age Ageing. 2018;48:16-31. doi:10.1093/ageing/afy169

9. Dodds RM, Syddall HE, Cooper R, Benzeval M, Deary IJ, Dennison EM, et al. Grip strength across the life course: normative data from twelve British studies. PloS one. 2014;9:e113637. doi:10.1371/journal.pone.0113637

10. Lauretani F, Russo CR, Bandinelli S, Bartali B, Cavazzini C, Di Iorio A, et al. Age-associated changes in skeletal muscles and their effect on mobility: an operational diagnosis of sarcopenia. Journal of Applied Physiology. 2003;95:1851–60.

11. Wang YC, Bohannon RW, Li X, Sindhu B, Kapellusch J. Hand-Grip Strength: Normative Reference Values and Equations for Individuals 18 to 85 Years of Age Residing in the United States. The Journal of orthopaedic and sports physical therapy. 2018;48:685-93. doi:10.2519/jospt.2018.7851

12. Albrecht BM, Stalling I, Bammann K. Sex- and age-specific normative values for handgrip strength and components of the Senior Fitness Test in community-dwelling older adults aged 65-75 years in Germany: results from the OUTDOOR ACTIVE study. BMC geriatrics. 2021;21:273. doi:10.1186/s12877-021-02188-9

13. Hofmann M, Halper B, Oesen S, Franzke B, Stuparits P, Tschan H, et al. Serum concentrations of insulin-like growth factor-1, members of the TGF-beta superfamily and follistatin do not reflect different stages of dynapenia and sarcopenia in elderly women. Experimental gerontology. 2015;64:35-45. doi:10.1016/j.exger.2015.02.008

14. Lima RM, de Oliveira RJ, Raposo R, Neri SGR, Gadelha AB. Stages of sarcopenia, bone mineral density, and the prevalence of osteoporosis in older women. Archives of osteoporosis. 2019;14:38. doi:10.1007/s11657-019-0591-4

15. Akpinar TS, Tayfur M, Tufan F, Sahinkaya T, Kose M, Ozsenel EB, et al. Uncomplicated diabetes does not accelerate age-related sarcopenia. The aging male : the official journal of the International Society for the Study of the Aging Male. 2014;17:205-10. doi:10.3109/13685538.2014.963040

16. Farinatti P, Paes L, Harris EA, Lopes GO, Borges JP. A Simple Model to Identify Risk of Sarcopenia and Physical Disability in HIV-Infected Patients. Journal of strength and conditioning research. 2017;31:2542-51. doi:10.1519/jsc.0000000000002070

17. Gadelha AB, Vainshelboim B, Ferreira AP, Neri SGR, Bottaro M, Lima RM. Stages of sarcopenia and the incidence of falls in older women: A prospective study. Arch Gerontol Geriatr. 2018;79:151-7. doi:10.1016/j.archger.2018.07.014

18. McGrath R, Hackney KJ, Ratamess NA, Vincent BM, Clark BC, Kraemer WJ. Absolute and Body Mass Index Normalized Handgrip Strength Percentiles by Gender, Ethnicity, and Hand Dominance in Americans. Advances in geriatric medicine and research. 2020;2: doi:10.20900/agmr20200005

19. Manini TM, Visser M, Won-Park S, Patel KV, Strotmeyer ES, Chen H, et al. Knee extension strength cutpoints for maintaining mobility. J Am Geriatr Soc. 2007;55:451-7. doi:10.1111/j.1532-5415.2007.01087.x

20. Foley KT, Owings TM, Pavol MJ, Grabiner MD. Maximum grip strength is not related to bone mineral density of the proximal femur in older adults. Calcified Tissue International. 1999;64:291–4.

21. Maranhao Neto GA, Oliveira AJ, Pedreiro RC, Pereira-Junior PP, Machado S, Marques Neto S, et al. Normalizing handgrip strength in older adults: An allometric approach. Archives of Gerontology and Geriatrics. 2017;70:230-4. doi:10.1016/j.archger.2017.02.007

22. Pua Y-H. Allometric analysis of physical performance measures in older adults. Physical therapy. 2006;86:1263–70.

23. Abdalla PP, Dos Santos Carvalho A, Dos Santos AP, Venturini ACR, Alves TC, Mota J, et al. Cut-off points of knee extension strength allometrically adjusted to identify sarcopenia risk in older adults: A cross-sectional study. Arch Gerontol Geriatr. 2020;89:104100. doi:10.1016/j.archger.2020.104100

24. Abdalla PP, Venturini ACR, Santos APD, Tasinafo M, Marini JAG, Alves TC, et al. Normalizing calf circumference to identify low skeletal muscle mass in older women: a cross-sectional study. Nutr Hosp. 2021;doi:10.20960/nh.03572

25. Davies MJ, Dalsky GP. Normalizing strength for body size differences in older adults. Medicine & Science in Sports & Exercise. 1997;29:713–7.

26. Bouchard DR, Beliaeff S, Dionne IJ, Brochu M. Fat Mass But Not Fat-Free Mass Is Related to Physical Capacity in Well-Functioning Older Individuals: Nutrition as a Determinant of Successful Aging (NuAge)—The Quebec Longitudinal Study. The Journals of Gerontology: Series A. 2007;62:1382-8. doi:10.1093/gerona/62.12.1382

27. Broadwin J, Goodman-Gruen D, Slymen D. Ability of Fat and Fat-Free Mass Percentages to Predict Functional Disability in Older Men and Women. Journal of the American Geriatrics Society. 2001;49:1641-5. doi:https://doi.org/10.1111/j.1532-5415.2001.49273.x

28. Enright PL. The Six-Minute Walk Test. Respiratory Care. 2003;48:783–5.

29. Binder EF, Yarasheski KE, Steger-May K, Sinacore DR, Brown M, Schechtman KB, et al. Effects of progressive resistance training on body composition in frail older adults: results of a randomized, controlled trial. The Journals of Gerontology: Series A. 2005;60:1425–31.

30. Cruz-Jentoft AJ, Landi F, Schneider SM, Zuniga C, Arai H, Boirie Y, et al. Prevalence of and interventions for sarcopenia in ageing adults: a systematic review. Report of the International Sarcopenia Initiative (EWGSOP and IWGS). Age and ageing. 2014;43:748-59. doi:10.1093/ageing/afu115

31. Finney GR, Minagar A, Heilman KM. Assessment of Mental Status. Neurologic clinics. 2016;34:1-16. doi:10.1016/j.ncl.2015.08.001

32. Icaza MC, Albala C. Projeto SABE. Minimental State Examination (MMSE) del estudio de dementia en Chile: análisis estatístico Brasília: OPAS. 1999;1–18.

33. WHO Expert Consultation. Appropriate body-mass index for Asian populations and its implications for policy and intervention strategies. Lancet (London, England). 2004;363:157-63. doi:10.1016/s0140-6736(03)15268-3

34. Segal NA, Torner JC, Yang M, Curtis JR, Felson DT, Nevitt MC. Muscle mass is more strongly related to hip bone mineral density than is quadriceps strength or lower activity level in adults over age 50 year. Journal of Clinical Densitometry. 2008;11:503-10. doi:10.1016/j.jocd.2008.03.001

35. Bailey BJ, Briars GL. Estimating the surface area of the human body. Statistics in medicine. 1996;15:1325–32.

36. Jelliffe DB, Jelliffe EP. The arm circumference as a public health index of protein-calorie malnutrition of early childhood. 20. Current conclusions. Journal of tropical pediatrics. 1969;15:253–60.

37. Heymsfield SB, McManus C, Smith J, Stevens V, Nixon DW. Anthropometric measurement of muscle mass: revised equations for calculating bone-free arm muscle area. Am J Clin Nutr. 1982;36:680–90.

38. George LB, Bruce RB, Baltej SM, Haran TS, Michael FS. Nutritional and metabolic assessment of the hospitalized patient. Journal of Parenteral and Enteral Nutrition. 1977;1:11-21. doi:10.1177/014860717700100101

39. Lean M, Han TS, Deurenberg P. Predicting body composition by densitometry from simple anthropometric measurements. The American journal of clinical nutrition. 1996;63:4–14.

40. Baumgartner RN, Koehler KM, Gallagher D, Romero L, Heymsfield SB, Ross RR, et al. Epidemiology of sarcopenia among the elderly in New Mexico. American journal of epidemiology. 1998;147:755–63.

41. Baumgartner RN, Heymsfield SB, Lichtman S, Wang J, Pierson RN, Jr. Body composition in elderly people: effect of criterion estimates on predictive equations. Am J Clin Nutr. 1991;53:1345-53. doi:10.1093/ajcn/53.6.1345

42. Lohman TG, Roche AF, Martorelli R. Anthropometric standardization reference manual. Champaing human kinects 1988;1:

43. Morley JE, Abbatecola AM, Argiles JM, Baracos V, Bauer J, Bhasin S, et al. Sarcopenia with limited mobility: an international consensus. J Am Med Dir Assoc. 2011;12:403-9. doi:10.1016/j.jamda.2011.04.014

44. Massy-Westropp NM, Gill TK, Taylor AW, Bohannon RW, Hill CL. Hand Grip Strength: age and gender stratified normative data in a population-based study. BMC Research Notes. 2011;4:1–5.

45. Lourenco R, Perez-Zepeda M, Gutierrez-Robledo L, Rodriguez Manas L, Garcia-Garcia F. Performance of the European Working Group on Sarcopenia in Older People algorithm in screening older adults for muscle mass assessment. Age and ageing. 2014;44:334-8. doi:10.1093/ageing/afu192

46. Alexandre S, Duarte YA, Santos JL, Wong R, Lebrao ML. Prevalence and associated factors of sarcopenia among elderly in Brazil: findings from the SABE study. The Journal of Nutrition, Health & Aging. 2014;18:284-90. doi:10.1007/s12603-013-0413-0

47. Brzycki M. Strength testing—predicting a one-rep max from reps-to-fatigue. Journal of Physical Education, Recreation & Dance. 1993;64:88–90.

48. Abdalla PP, Carvalho AdS, dos Santos AP, Venturini ACR, Alves TC, Mota J, et al. One-repetition submaximal protocol to measure knee extensor muscle strength among older adults with and without sarcopenia: a validation study. BMC Sports Science, Medicine and Rehabilitation. 2020;12:29. doi:10.1186/s13102-020-00178-9

49. Matsudo S, Araújo T, Matsudo V, Andrade D, Andrade E, Oliveira LC, et al. Questionário internacional De atividade física (ipaq): estupo De validade e reprodutibilidade No Brasil. Revista Brasileira de Atividade Física & Saúde. 2012;6:5–18.

50. Myers R. Classical and modern regression with applications. Boston: PWS and Kent Publishing Company. Inc; 1990.

51. Mukaka MM. Statistics corner: A guide to appropriate use of correlation coefficient in medical research. Malawi Med J. 2012;24:69–71.

52. Schisterman EF, Perkins NJ, Liu A, Bondell H. Optimal cut-point and its corresponding Youden Index to discriminate individuals using pooled blood samples. Epidemiology. 2005;16:73–81.

53. Hosmer D, Lemeshow S. Applied logistic regression. 2 ed. Nova Jersey, EUA: John Wiley & Sons; 2000.

54. Cawthon PM, Manini T, Patel SM, Newman A, Travison T, Kiel DP, et al. Putative Cut-Points in Sarcopenia Components and Incident Adverse Health Outcomes: An SDOC Analysis. Journal of the American Geriatrics Society. 2020;68:1429-37. doi:https://doi.org/10.1111/jgs.16517

55. Alley DE, Shardell MD, Peters KW, McLean RR, Dam T-TL, Kenny AM, et al. Grip Strength Cutpoints for the Identification of Clinically Relevant Weakness. The Journals of Gerontology: Series A. 2014;69:559-66. doi:10.1093/gerona/glu011

56. Owings TM, Pavol MJ, Grabiner MD. Lower extremity muscle strength does not independently predict proximal femur bone mineral density in healthy older adults. Bone. 2002;30:515–20.

57. Ramírez-Vélez R, Sáez de Asteasu ML, Martínez-Velilla N, Zambom-Ferraresi F, García-Hermoso A, Izquierdo M. Handgrip Strength as a Complementary Test for Mobility Limitations Assessment in Acutely Hospitalized Oldest Old. Rejuvenation Res. 2021;doi:10.1089/rej.2020.2344

58. Jaric S. Role of body size in the relation between muscle strength and movement performance. Exercise and Sport Sciences Reviews. 2003;31:8–12.

